# Effectiveness of Stress Management to Reduce Stress Eating for Women: A Systematic Review and Meta-analysis of Intervention Studies

**DOI:** 10.64898/2026.06.11.26355007

**Authors:** Vanessa V. Volpe, Abbey N. Collins, Elizabeth M. Davis, Oreoluwa O. Badejoh, MarQia Allen, Melissa C. Holland, Julia M. Ross, Abby Braden, Keri F. Kirk, Emily C. Hector

## Abstract

**Objective:** This systematic review and meta-analysis examined 1) the effects of stress management interventions on changes in stress eating for women, and 2) the longevity of these effects, by summarizing and assessing evidence from controlled and non-equivalent pretest-posttest intervention studies.

**Method:** Five databases (PsycINFO, PubMed, Medline, Web of Science, CINAHL), existing sources, and grey literature were searched (February - June 2025). Studies that assessed stress eating or emotional eating, included a stress management intervention, and comprised at least 70% women were included. The primary outcome was reduction in stress eating. Data were pooled in meta-analyses using multi-level random-effects models and subset by follow-up period. Risk of bias was assessed via funnel plots and sensitivity analyses.

**Results:** Sixty studies with 119 effect size estimates were included in the primary analysis. Pooled estimates indicated that stress management interventions significantly reduced stress eating (Hedges’ *g* = -0.4174, p < 0.001), with pre-post designs having larger effects than controlled trials. Subgroup analyses of follow-up periods found small effects in the short-term (before 3 months; Hedges’ *g* = -0.4202, *p* < 0.0001) and moderate effects for mid-term (3-6 months; Hedges’ *g* = -0.5886, *p* < 0.0001). Effects beyond 6 months were small and nonsignificant (Hedges’ *g* = -0.4370, *p* = 0.0660).

**Conclusion and Relevance:** Stress management interventions appear to be effective for reducing stress eating for women, suggesting the potential to incorporate stress management in interventions targeting obesity. Effects may be only sustained 6 months post-intervention, suggesting the need for strategies to bolster long-term effectiveness.

## 1. Introduction

Stress eating, defined in this investigation as eating in response to feelings of stress, is a central health behavior related to cardiometabolic risk for women [1,2]. Stress eating may involve loss of control of eating in response to negative emotional states (i.e., emotional eating [3]) and the psychosocial stress experience (i.e., stress eating [4]). A majority of overweight and obese women enrolled in weight management programs struggle with stress eating [5]. However, stress eating is rarely the target of lifestyle interventions for weight management compared to behaviors such as diet, physical activity, and self-weighing. Therefore, reducing stress eating for women remains a top priority for the effectiveness of current and future obesity interventions and treatment for this population.

Stress management interventions may substantially reduce barriers to weight management for women at high health risk by reducing stress eating, but we do not yet have sufficient evidence of their effectiveness and longevity. Several systematic reviews and meta-analyses have been conducted that highlight the importance of research in this area. For example, meta-analyses have found that more stress and negative emotions are linked to eating in correlational studies [6–8], that stress eating interventions improve stress eating and weight management outcomes [9–10], and that some stress management interventions improve weight management outcomes [11–12]. However, no current meta-analysis examines the effectiveness and longevity of stress management interventions to improve stress eating outcomes among women. One review established a connection between greater emotional eating and body mass index and suggested that interventions that leverage cognitive-behavioral and mindfulness-based approaches are somewhat effective with small effect sizes in the short-term (i.e., immediately post-intervention)[13]. However, this review used description of effect sizes in the existing literature to generate this conclusion, rather than conducting a meta-analysis of effect sizes, leaving questions about study heterogeneity and the precision of estimates of effects. It also focused on behavioral weight loss programs, rather than considering all potential stress management interventions that may help women with obesity reduce stress eating in non-clinical settings. Another promising meta-analysis considered mindfulness-based interventions for obesogenic eating behaviors, finding a small non-significant overall effect for stress eating behaviors [14]. However, this investigation focused only on one type of stress intervention and examined effectiveness at only one post-intervention timepoint. This limits our ability to understand 1) the range of stress management interventions effective for stress eating and 2) the longevity of any effects. The current meta-analysis aims to address both of these gaps to better inform the development and refinement of stress management interventions to reduce stress eating for women across a range of settings, thereby contributing to the prevention and better management of overweight and obesity.

### 1.1. Obesity, Women, and Stress Eating

Obesity is a central driver of cardiometabolic disease [15–16] and all-cause mortality [17] and costs the United States (US) billions per year [18]. Women have a higher risk of cardiometabolic disease than is often known or acknowledged, leading to gaps in treatment and diagnosis and resulting in higher rates of cardiometabolic mortality [19]. For instance, women experience delays in receiving care, poorer outcomes when treated by male physicians, and lower rates of guideline-based medical therapy than men [20–22]. Therefore, developing strategies to mitigate obesity risk specifically for women is necessary.

One contributor to increased obesity risk is stress exposure [23], because a common response to stress is engagement in stress eating. Stress eating is a way to regulate mood at the physiological level and serves as a means of psychological escape from distressing thoughts [24]. Women are particularly impacted by stress and report high levels of stress eating [25–27].Therefore, intervening upon stress eating by reducing stress for women could be a pathway to better cardiometabolic health by reducing obesity risk. In this way, interventions that reduce stress eating for women are a priority for behavioral medicine to reduce the disease burden of obesity.

Several prior investigations of stress eating interventions suggest that reducing stress eating facilitates better weight management, especially among individuals with overweight or obesity [5,9,12–13]. Yet it remains unclear if stress interventions broadly - including but not limited to stress *eating* interventions - can reduce stress eating. This is an important question because broader stress management interventions, if found effective for stress eating, may provide an additional general benefit to a variety of clinical outcomes. It may be easier to add a stress management component to a wider range of existing interventions that could reduce stress eating and improve obesity risk, rather than considering only the need for a targeted or full stress eating intervention in all contexts. Therefore, in this systematic review and meta-analysis, we aimed to take a broader perspective to consider the benefits of stress management interventions writ large. We focus on stress management interventions and their impact on stress eating, which in some cases are discrete stress eating interventions or behavioral weight loss interventions, and in other cases are portable stress management interventions or general lifestyle interventions with stress management components. This broader perspective considers the degree to which these interventions with at least one active component of stress management strategy are effective in a variety of contexts and populations - whether eating is a primary or secondary target. Casting this wider net allows us to consider the promise of such interventions for both stress eating prevention in non-clinical samples and occurrence among those enrolled in existing biobehavioral interventions with overweight or obesity, responsibly addressing barriers to obesity treatment across the spectrum of disease risk.

### 1.2. Stress Management Interventions and Stress Eating

Stress management involves techniques from various modalities that target the reduction of, management of, or coping with stress. The components include strategies generally used to manage or cope with experienced stress, such as psychoeducation, problem-solving, coping or coping skill(s), cognitive reframing, cognitive restructuring, skills building, skill(s) acquisition, cognitive reappraisal, mindfulness, compassion, relaxation, biofeedback, acceptance, emotion regulation, and psychotherapeutic modalities. Specifically, modalities like Acceptance and Commitment Therapy (ACT), Dialectical Behavioral Therapy (DBT), Cognitive Behavioral Therapy (CBT), and stress inoculation therapy are all used to address stress management in various forms. Although there is considerable heterogeneity among interventions that use these strategies, generally, such interventions may work in one of three interrelated ways: 1) by examining and reshaping core cognitions that may be contributing to stress (e.g., cognitive restructuring), 2) by learning and practicing new affect regulation strategies that do not involve food (e.g., mindfulness), and 3) by building distress tolerance and emotional approach in stressful situations. For example, CBT aims to reframe maladaptive thought processes, thereby reducing stress from rumination or catastrophizing [28]. Stress eating interventions focused on emotion regulation and distress tolerance often try to disrupt the cycle of using food to distract from and reduce negative affect [29–30]. Therefore, stress management interventions that promote non-eating coping strategies to regulate emotions could reduce stress eating.

Many stress management interventions target mental health symptoms or cardiovascular reactivity, with relatively fewer focused on weight-related outcomes and fewer still that explicitly aim to reduce stress eating. The effectiveness of stress management has been considered in previous systematic reviews or meta-analyses focused on weight [11,31], cardiovascular disease risk [32–33], and eating disorders [34–35]. However, no current meta-analysis examines the role of stress management on stress-related non-clinical eating behaviors (i.e., eating behaviors not associated with an eating disorder diagnosis), which are intermediary health behaviors that govern cardiometabolic risk. Our work aims to fill that gap, thereby directly contributing to the design of tailored interventions that target stress-related non-clinical eating behaviors for women.

### 1.3. Current Study Aims

The aim of this review and meta-analysis is to examine the effectiveness of stress management interventions on the reduction of stress eating and the longevity (or lack thereof) of these reductions for women. This review quantifies the strength of unique stress management interventions writ large on stress eating for women (Aim 1), clarifying their potential for behavioral medicine and obesity science. Furthermore, the present review seeks to provide insights about potential length of any observed effects (Aim 2), which is an important insight for existing weight management or loss interventions hoping to consider increased effectiveness by incorporating a stress management component. Taken together, this investigation aims to inform research on future interventions that are maximally beneficial and long-lasting in reducing health risks from obesity for women.

## 2. Materials & Methods

Online replication code and supplementary materials are available at https://github.com/emdavis7/stress_eating_meta_analysis.

### 2.1. Literature Search

The current systematic review sought to identify intervention studies (i.e., controlled trials, non-equivalent groups designs as in pre-post test no control group, basic experimental studies with humans) that tested the effects of stress management interventions on stress eating. We followed design and reporting guidelines from the Preferred Reporting Items for Systematic Reviews and Meta-Analyses (PRISMA) Statement [36] (see Supplemental Material 1). Our larger systematic review and meta-analysis project ^1^ was pre-registered on April 21, 2024 via the International Prospective Register of Systematic Reviews (PROSPERO) entitled “Effectiveness of Stress Management to Reduce Stress Eating for Women: A Systematic Review and Meta-analysis” (CRD42024534775). See Figure 1 for our search and screening process.

**Figure 1.**
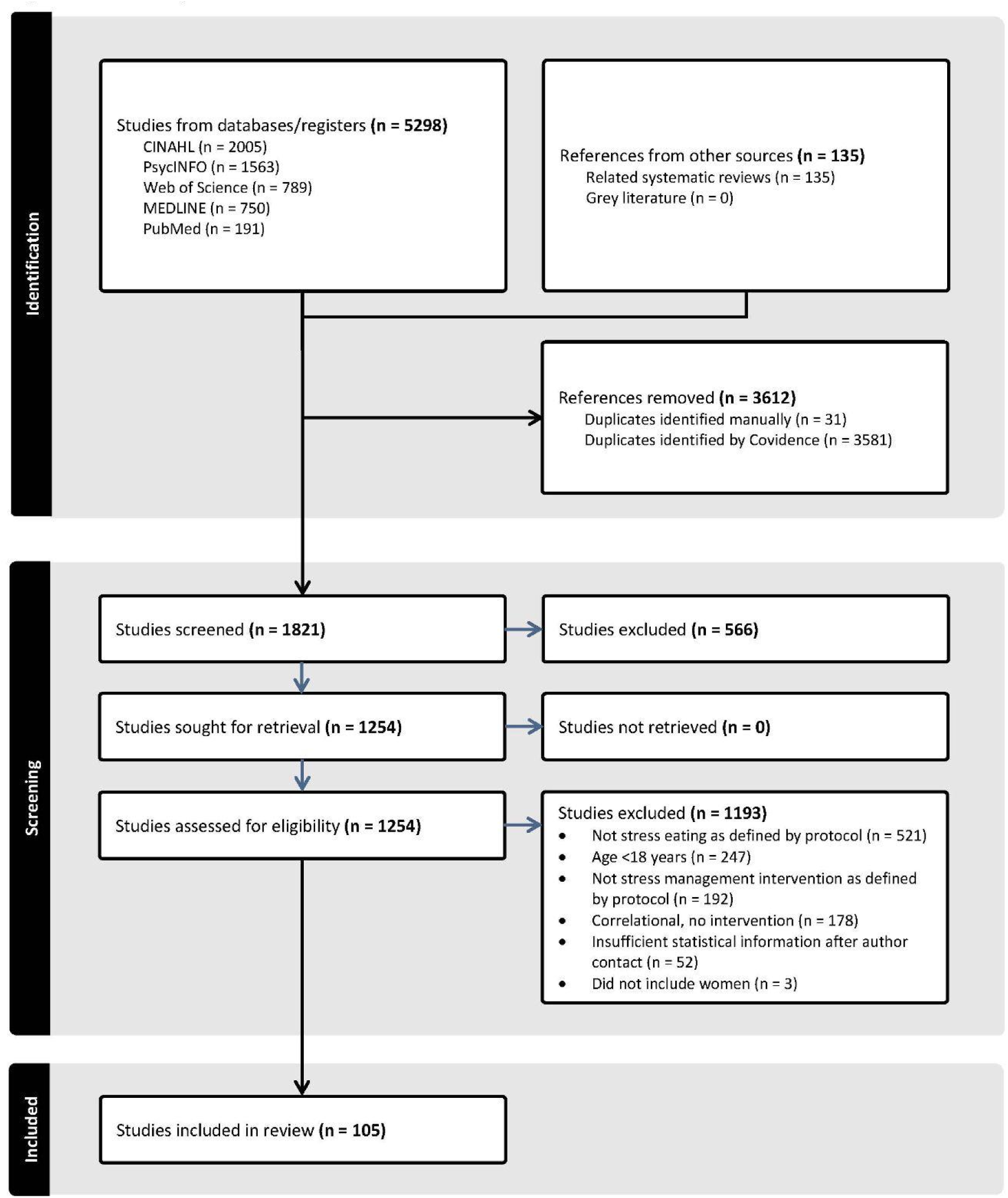
PRISMA Diagram. *Note:* During February 2025 when we were conducting our search, the database platform EBSCOHost updated its interface, resulting in significantly more references downloaded into the bibliographic data file than included in the search itself. As the timeline for resolving this known platform issue was uncertain, we opted to continue with the screening process. This likely resulted in the high number of initial studies screened out that are reflected in the PRISMA diagram.

### 2.2. Eligibility Criteria

We included studies that met the following criteria: 1) written in English, 2) sample is >= 70% women, 3) sample includes adults age >= 18, 4) study examines at least one stress eating behavior as a primary outcome, 5) study includes an intervention with at least one active component being a stress management strategy, 6) study includes one or more quantitative results specific to the effect of stress management on stress eating. Studies were excluded if they were case studies, single-case experiments, theoretical or conceptual papers, systematic reviews or meta-analyses, book reviews or chapters, abstracts or conference proceedings, theses or dissertations, or treatment guidelines or manuals. Non-human animal studies were also excluded. Proof-of-concept papers and pilot studies were included.

From a review of the existing literature, we generated an exhaustive list of non-clinically-disordered stress eating behaviors, which included: stress eating, stress-induced eating, stress-related eating, eating to cope, eating in response to negative emotions, and emotional overeating. Studies that only measured eating disorder diagnosis or severity, nutrition, caloric intake, or metabolic expenditure were excluded. Studies with samples of only those with eating disorders were excluded. Studies that used clinical measures of eating disorder symptomatology (e.g., Eating Disorders Scale, Eating Disorder Examination Questionnaire) or did not explicitly include constructs related to stress eating (e.g., Eating Attitudes Test) were excluded, as this review focused on stress eating as a behavior among those without diagnosed eating disorders.

From a review of common foci of stress management interventions in health psychology, we generated a list of relevant stress management strategies. Strategies included: psychoeducation, problem-solving, coping, cognitive reframing, cognitive restructuring, stress management skills building, stress management skills acquisition, cognitive reappraisal, mindfulness, mindful eating or intuitive eating, compassion, relaxation, biofeedback, acceptance, emotion regulation, and psychotherapeutic modalities (ACT, DBT, CBT, cognitive therapy, attention and interpretation therapy, stress inoculation) if they explicitly target stress. Studies that examined stress management strategies that were purely pharmacological (e.g., psychedelic-assisted therapy, medications) were excluded.

### 2.3. Search Strategy

Studies were identified by searching five electronic databases (PsycINFO, PubMed, Medline, Web of Science, CINAHL) using a Boolean search strategy (see Supplemental Material 2). Searches were conducted in February 2025. We also reviewed reference lists from relevant systematic reviews and meta-analyses and articles authored by identified experts in this area, and clinical trials databases (clinicaltrials.gov, Cochrane Central Trials Register) in May and June 2025. To minimize publication bias, we sent requests for unpublished data on this topic via relevant professional listservs (e.g., Society for Health Psychology, Society for Behavioral Medicine) in June 2025.

### 2.4. Data Screening and Extraction

Screening articles for potential inclusion proceeded in several steps and was facilitated using the systematic review and meta-analysis tool Covidence Systematic Review Software [37]. Studies from the search were imported into Covidence. Duplicates were automatically identified by the tool and were also manually identified by coders as they progressed through screening. All records were first screened based on title and abstract by four trained coders. Next, these coders retrieved and reviewed full-text manuscripts of potentially relevant records for inclusion based on the inclusion/exclusion criteria detailed above. Screening discrepancies were resolved by the principal investigator.

After screening, relevant study information (e.g., publication year), sample characteristics (e.g., percentage of women, mean age), design (e.g., controlled trials vs. pretest-posttest studies, type of control group), intervention details (e.g., delivery modality), and intervention components (e.g., stress management strategies) were extracted by two trained coders using detailed coding forms. Extraction of final study information was verified by the principal investigator. Studies were categorized based on the type of stress management intervention and type of stress eating outcome. Studies that included multiple stress management interventions and/or eating outcomes were included within each relevant category.

For the meta-analytic portion of the study, three trained independent coders completed the extraction of data for the calculation of effect sizes for all relevant stress eating outcomes defined in the inclusion criteria. Discrepancies between coders were resolved through discussion, corrected, and finalized. Effect size data were verified by the principal investigator. Pre-intervention and all post-intervention time points with stress eating data on any measure that were available were extracted. When studies included multiple comparator interventions or did not include a control group, we extracted pre-post scores for all groups that met our stress intervention criteria. In cases where intervention groups were separated by other considerations not of interest to the current meta-analysis (e.g., accompanying medical dosage), we collapsed groups accordingly and treated the study as a pre-post investigation. For studies that included greater than 0% and less than 70% of women, authors were contacted to provide statistics only for the women in the sample. Authors were also contacted if methodological details needed clarification or effect size estimates were missing, in an attempt to include as many studies as possible. Sixty-one authors were contacted for additional statistical information (e.g., missing means and standard deviations, pre-test post-test correlations) and 20 (32.79%) provided the information requested within three weeks.

To handle missing summary statistics, we converted between relevant statistical estimates when the availability of statistical information made it possible (e.g., mathematically calculating a standard deviation from a reported standard error). For non-equivalent groups designs that required some estimate of correlation between timepoints (i.e., pre-posttest no control group) but for which such information was unavailable after three weeks of author contact, in a number of cases (*k* = 18; *n* = 37, 30.83% of effect sizes), we input .5 as an estimate. If an effect size could not be computed due to missing summary statistics and the author was non-responsive to follow-up after three weeks of contact, that effect size was excluded.

### 2.5. Meta-analysis

For controlled trials, summary effect sizes were calculated as the standardized mean difference between the stress management intervention and comparison groups, controlling for baseline. To calculate effect sizes for each outcome in the trial, we used the mean, standard deviation, and the sample size in the intervention and control groups at each data assessment post-intervention. To calculate effect sizes for each outcome for non-equivalent groups designs, we used mean differences or standardized mean differences, standard deviations of the differences or pooled standard deviations, and correlations between assessment timepoints. For both controlled trials and non-equivalent groups designs, we estimated the effect sizes with Hedge’s *g* to improve estimation for studies with small sample sizes. Hedge’s *g* estimate magnitudes were interpreted as small = 0.20, medium = 0.50, or large = 0.80. Negative effect sizes indicated that stress management interventions improved (i.e., reduced) stress eating. Among outcomes where positive effect sizes indicated an improvement in stress eating (e.g., an outcome variable was “ability to stop” emotionally eating such that a higher score would indicate a beneficial reduction in emotional eating behavior), the effect size sign was reversed.

Included studies could report multiple interventions, multiple outcomes, and multiple timepoints. We accounted for the correlation of effect sizes in our analysis using nested random effects. Correlation across time was modeled with a continuous-time autoregressive structure and was nested within each outcome measured. For studies with multiple outcomes and interventions, outcomes were modeled as nested within interventions, which were nested within study, for estimation of variance (See Supplemental Material 3). Our primary analysis was a pooled effect size with all time points and was conducted with restricted maximum likelihood estimation. Effect sizes and 95% confidence intervals were estimated using cluster-robust standard errors and the *t* distribution, which are appropriate for dependent effect sizes [38]. In this case, our cluster-robust estimation supplants the need for the traditional Knapp-Hartung estimate. We display these confidence intervals and the pooled effect with a forest plot.

We measured the degree of heterogeneity among studies with the *Q* statistic; a significant *Q* indicates that the effects are not homogeneous. We also calculated the *I^2^* value, which shows the percent of variation in effects attributable to heterogeneity [39]. An *I^2^*value of 25% indicates low heterogeneity, 50% indicates moderate heterogeneity, and 75% and higher indicates high heterogeneity [40]. We additionally reported the percent of variation in effects that is attributable to between-study variability, rather than between-intervention or between-outcome variability. We examined the influence of individual studies by recording the pooled effect size, *p*-value, and *I^2^* when effects from the study were excluded.

To investigate the heterogeneity in the effect sizes, we consider the design of the study (controlled trial vs non-equivalent groups) and the length of the intervention as moderators, in two separate analyses. We additionally consider the durability of interventions over time. We grouped effects into those measured mid-intervention (prior to the end of the intervention), short-term effects (measured less than 3 months post-intervention), mid-term effects (measured at 3 or 4 months post-intervention), and long-term effects (measured from 6 months to a year post-intervention). We analyzed effects in these groups separately, using the same structure of random effects nested within study, intervention, and outcome as described above. Within these groups, the design of the study is confounded with the timing of the measurement ^2^. We therefore did not include the design of the study as a moderator in the subgroup analysis. All analyses were conducted in R (version 4.5.1), using the metafor package (version 4.8-0) [41].

### 2.6. Risk of Bias

To assess the risk of publication bias in our effect sizes, we first constructed a funnel plot to visually check for asymmetry. Funnel plots for moderator analyses, subgroup analyses, and sensitivity analyses are shown in Supplemental Figures. We further conducted the Begg and Mazumdar rank test for funnel plot asymmetry and Egger’s regression asymmetry test. To assess the sensitivity of our results to inputting a 0.5 for correlation between time points, we reanalyzed the data excluding the pre-post studies for which this correlation was missing. We also reanalyzed the data including one study for which the effect direction conflicted with the reported pre- and post-treatment means in the published paper. We conducted two additional subgroup analyses, changing the follow-up period grouping patterns. In the first, we assessed the immediate post-intervention effect as its own category and re-assessed the short-term effect without these observations. In the second, we re-assessed the short-term effect but considered effects measured at 2 months post-intervention within the “medium-term” category.

## 3. Results

After duplicates were removed, a total of 1821 unique articles were retrieved via our search strategy (see Figure 1 for PRISMA diagram, see Supplemental Material 4 for list of included studies). Next, 566 were excluded at the title and abstract review level, with the most common reasons being that they were irrelevant. ^3^ Others were not written in English, were not peer-reviewed journal articles (e.g., dissertations, conference abstracts), or did not include quantitative data (e.g., existing reviews, conceptual manuscripts, qualitative studies). Of the 1254 studies assessed for eligibility at the full text level, a majority (*k* = 1193) were excluded for reasons such as lack of examination of key constructs as defined by the protocol, sample mismatch (i.e., pediatric populations, samples comprised of only men), correlational studies that did not examine an intervention, and insufficient provision of statistical information after author contact. A final 61 studies were considered for inclusion in this review, generating a total of 120 effect sizes. Table 1 displays the nesting structure for the included studies.

**Table 1.** Description of Study Nesting.

| **Intervention** | **Outcome** | **Time point** | **Number of Studies** |
| --- | --- | --- | --- |
| One | One | One | 7 |
|  |  | Multiple | 4 |
|  | Multiple | One | 29 |
|  |  | Multiple | 16 |
| Multiple | One | One | 2 |
|  |  | Multiple | 0 |
|  | Multiple | One | 1 |
|  |  | Multiple | 2 |

### 3.1. Study characteristics

Among the 61 included studies, 29 (47.5%) were conducted in the United States (US). Most investigations conducted outside of the US were conducted in Canada (*k* = 7), Brazil (*k* = 6), the Netherlands (*k* = 5), and Portugal (*k* = 4). The combined sample size from the included studies was 3411 (*M* = 55.91, *SD* = 97.40; range 3 to 743 in individual studies). Of the 61 studies, a majority (*k* = 32; 52.5%) used exclusively female participants and the average percentage of women included across studies was 93.63% (*SD* = 8.95%; range 70% - 100%). Mean age for the total sample was 41.52 years (*SD* = 9.87 years; range 21 to 59 years), with six studies not reporting mean age. On average, studies included young adults (83.33% included some young adults) and middle adults (90% included some middle adults), but fewer included older adults (33.33% included some older adults). Among studies conducted in the US in which race/ethnicity of participants was reported, White participants were the most common (*M* = 69.17% per study; *SD* = 23.33%) and 67.90% of studies included samples of at least 70% of White individuals. Four studies conducted in the US did not report racial/ethnic composition.

On average, study samples consisted of 91.70% (*SD* = 26.35%; range 0% to 100%) of participants with overweight or obesity (i.e., with a body mass index (BMI) over 25 kg/m^2^). Although BMI was not reported in 19 studies, 3 studies (7.14% of studies reporting BMI) included only healthy weight individuals (defined as a BMI from 18.5 to 24.9 kg/m^2^). Most studies reporting BMI (88.10%) included only participants with overweight or obesity.

Under our conceptual umbrella of stress eating, many studies examined emotional eating outcomes (*k* = 59; 96.72%), while the remainder examined stress eating outcomes (*k* = 2; 3.28%). These studies utilized self-reports of eating, with emotional eating largely measured using the Emotional Eating Questionnaire, Emotional Eating Scale, or emotional eating subscales of the Dutch Eating Behavior Questionnaire or Three-Factor Eating Questionnaire. Stress eating was measured using the Salzburg Stress Eating Scale or subscales of the Regulation of Eating Behavior Scale.

### 3.2. Intervention characteristics

About half of included studies were controlled trials with intervention groups (50.8%, *k* = 31) and the other half were pre-post intervention studies without control groups (49.2%, *k* = 30). Among those studies utilizing intervention groups, the majority (*k* = 28; 90.32%) randomly assigned individuals or clusters to groups. Approximately 61% (*k* = 19) used parallel group designs while the remainder (*k* = 12, 38.71%) used crossover designs. Most control conditions were waitlist (*k* = 10, 32.3%) or active controls (*k* = 10, 32.3%), while others were negative control groups (*k* = 8, 25.8%) or treatment as usual controls (*k* = 3, 9.7%).

Interventions employed a range of stress management strategies as active components. The most common was acceptance-based approaches including ACT psychotherapeutic modalities, acceptance, self-compassion, intuitive eating, mindfulness, or mindful eating (*k* = 31, 50.5%). Approximately eight percent of studies (*k* = 5) used behavioral approaches such as BAT, biofeedback, relaxation strategies like meditation, imagery, breathwork, or visualization. Approximately five percent used cognitive approaches such as problem solving, motivational interviewing, cognitive framing, restructuring, reappraisal, and cognitive therapy (*k* = 3), and approximately three percent used self-regulation strategies (*k* = 2). Only one study (1.6%) used stress psychoeducation. About a third of studies (*k* = 19, 31.1%) used a combination of one or more cognitive, behavioral, acceptance-based, self-regulation, or psychoeducation strategies, including but not limited to psychotherapeutic modalities like CBT (i.e., both cognitive and behavioral) and DBT (i.e., both self-regulation and acceptance).

Group-based interventions were most common (60.3% of interventions), with 19% focused on the individual, and 20.6% including both group and individual components. The majority (46%) of interventions were delivered in-person by trained facilitators or personnel (e.g., dietician, psychologist), followed by digital or print self-initiated content or exercises (11.1%) and digital or phone-based interventions delivered by trained facilitators or personnel (3.2%). Twenty-seven percent of interventions used both in-person and self-initiated delivery modes, 7.9% used both self-initiated and digital or phone-based delivery, 1.6% used both in-person and digital or phone-based delivery, and 3.2% used all three delivery modes.

Length of follow-up assessments of stress eating outcomes also varied for effect sizes included in our meta-analyses. We included nine mid-intervention effect sizes. Most studies reported results for immediate post-intervention effects (58%). Short-term follow-ups of 1 week (2.3%), 2 weeks (2.3%), 1 month (2.3%), or 2 month (1.1%) post-intervention were reported by fewer studies. In terms of longer follow-ups, three month (13.6%) and six month (6.8%) follow-ups were more common, and four month (2.3%) and one year (2.3%) follow-ups were less common. Mean intervention length was 14.20 weeks (*SD* = 15.72 weeks, range = .14 - 78.27 weeks). Retention from study enrollment (i.e., pre-posttest studies) or randomization (i.e., controlled trials) to final follow-up among included studies was 79.30%, on average (*SD* = 17.45, range = 25.41% - 100.00%).

### 3.3. Intervention pooled effects

The heterogeneity among the 119 effects, including mid-intervention measurements, is high with an *I*^2^ of 81.0% and *Q =* 448.5 (*p* < 0.0001) ^4^. The between-study variation accounts for 67.6% of the variability in the data. The funnel plot is approximately symmetric (see Figure 2). The Begg and Mazumdar rank test and Egger’s regression intercept test are both non-significant, with *p*-values of 0.1482 and 0.4721, respectively; there is no significant evidence of publication bias. Overall, interventions were associated with a small reduction in stress eating (*g* = -0.4174, *p* < 0.0001; 95% CI: -0.5334 to -0.3015). Supplemental Figure 1 displays the forest plot of these effects for all studies. We re-ran this analysis, leaving one study out each time, and determined that no studies were influential or disproportionately contributing to heterogeneity; the minimum *I*^2^ was 77.7%, and effects remained small (*g* from -0.4377 to -0.3972) and statistically significant for all runs.

**Figure 2.**
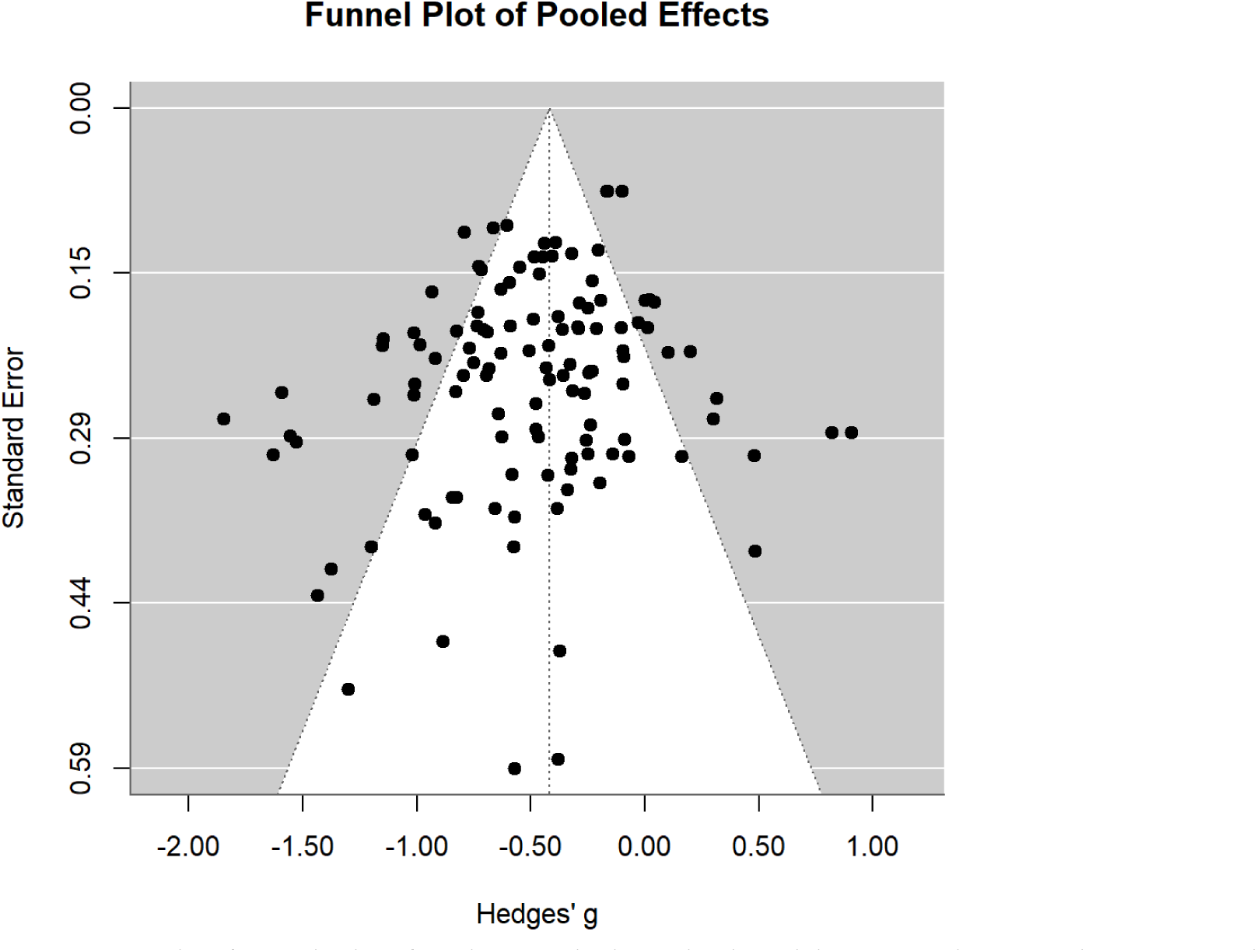
The funnel plot for the pooled analysis without moderators is symmetric.

Including the design of the study (i.e., controlled trials, pre-post designs) as a moderator decreases the heterogeneity, yielding an overall *I*^2^ of 74.9% (*Q =* 308.0, *p <*0.0001). With the moderator, 60% of the variability is attributable to variation among studies. Controlled trials have a significant and small effect size of -0.2058 (*p* = 0.003, 95% CI: -0.3370 to -0.0747). The effect size for pre-post designs is larger in magnitude than controlled trials by 0.4145 (*p* < 0.0001, 95% CI: -0.6150 to -0.2141). No studies were influential when re-running the analysis with one study left out; the minimum *I*^2^ was 70.7%, the effect size in controlled trials remained small and significant (*g* from -0.2378 to -0.1885), and the change in effect size for pre-post designs was small-to-moderate and significant in all cases (*g* from -0.4403 to -0.3765).

We additionally assessed the potential role of intervention length (in weeks) in the effect size. With the intervention length as a moderator, the heterogeneity remained high (*I*^2^ = 81.0%, *Q =* 441.3, *p <*0.0001). The length of the intervention was not significantly related to the effect size (*g = -*0.0017, *p* = 0.5098; 95% CI: -0.0076 to 0.0043). No single study was influential; when re-analyzing the data with one study excluded, the minimum *I*^2^ was 77.9% and all moderator effects were near zero and non-significant (*g* from -0.0028 to -0.0009).

### 3.4. Intervention effects by time

In the subgroup of 9 effects measured mid-intervention (see Figure 3), the effect size is small and significant (*g* = -0.3420, *p* =0.0484; 95% CI: -0.6800 to -0.0040). The effects display high heterogeneity (*I*^2^ = 82.7%, *Q* = 52.7, *p* < 0.0001), and there is not significant evidence of publication bias (Begg and Mazumdar *p =* 0.6122; Egger’s regression intercept *p* = 0.414).

**Figure 3.**
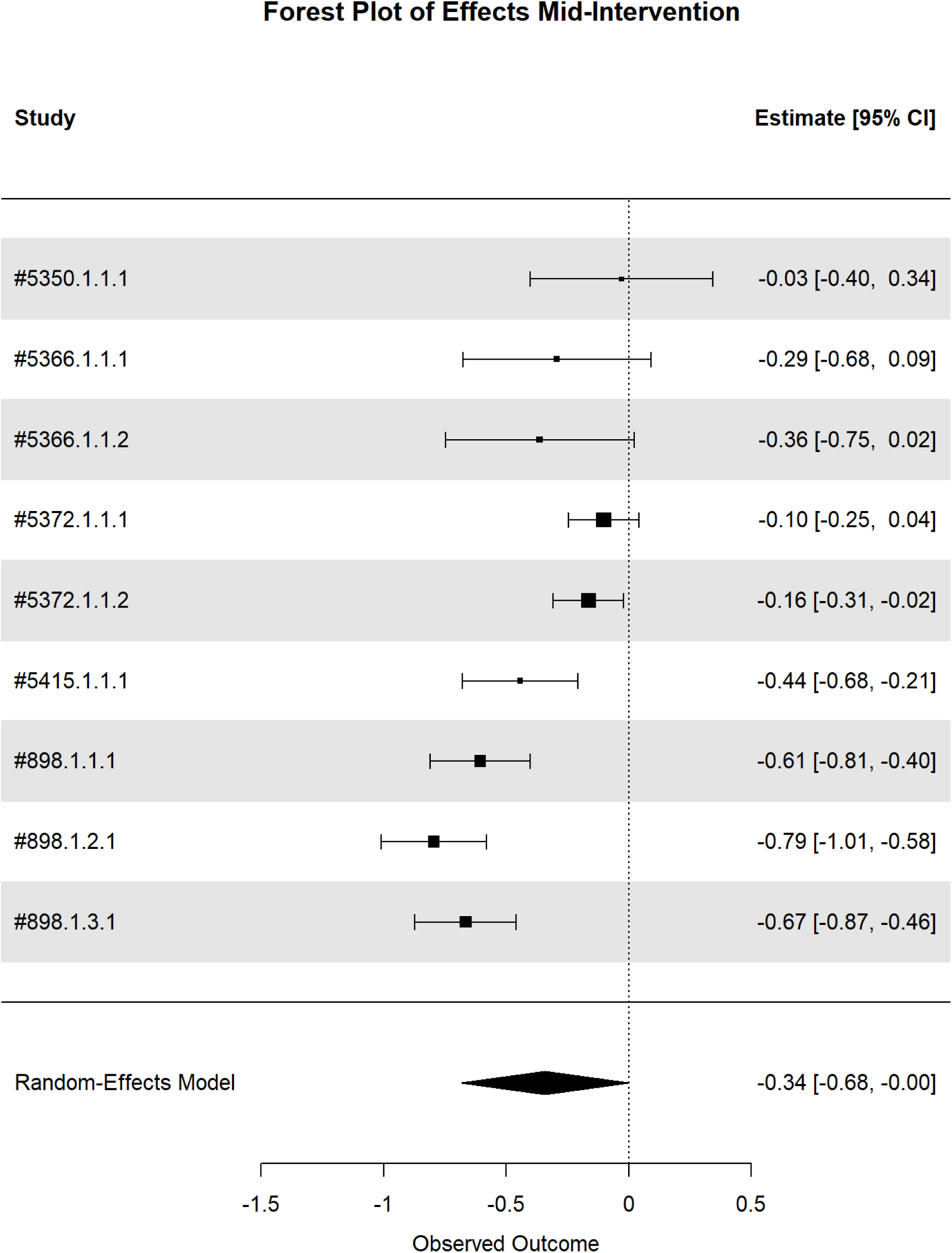
Forest plot for mid-intervention study follow-up timepoints.

For the subgroup of 80 effects measured less than 3 months post-intervention (see Figure 4), interventions were associated with a small-to-moderate, significant reduction in stress eating (*g* = -0.4202, *p* < 0.0001; 95% CI: -0.5446 to -0.2961). The effects display high heterogeneity (*I*^2^ = 78.5%, *Q* = 270.8, *p* < 0.0001), and there is not significant evidence of publication bias (Begg and Mazumdar *p =* 0.2028; Egger’s regression intercept *p* = 0.2688).

**Figure 4.**
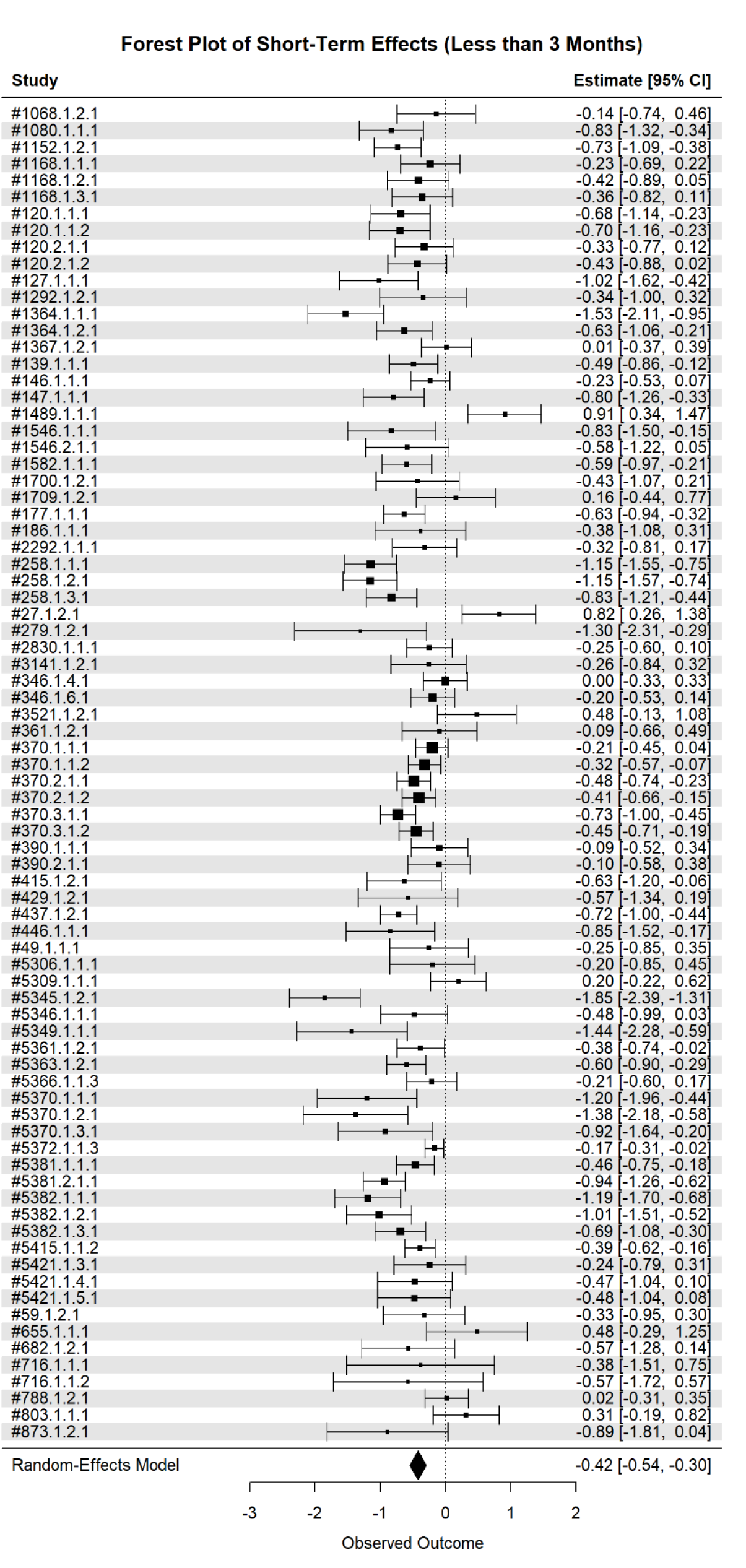
Forest plot for shorter-term post-intervention study follow-up timepoints.

The 17 medium-term effects, measured from 3 months to less than 6 months post-intervention (see Figure 5), have a moderate and significant effect (*g* = -0.5886, *p <* 0.0001; 95% CI: -0.8004 to -0.3769). The heterogeneity within this group is moderate, accounting for 48.3% of the variability (*Q* = 31.3, *p* < 0.0122). There was not substantial evidence of publication bias in this subgroup; the Begg and Mazumdar rank test and Egger’s regression intercept test were non-significant (*p* = 0.5976 and *p* = 0.412, respectively).

**Figure 5.**
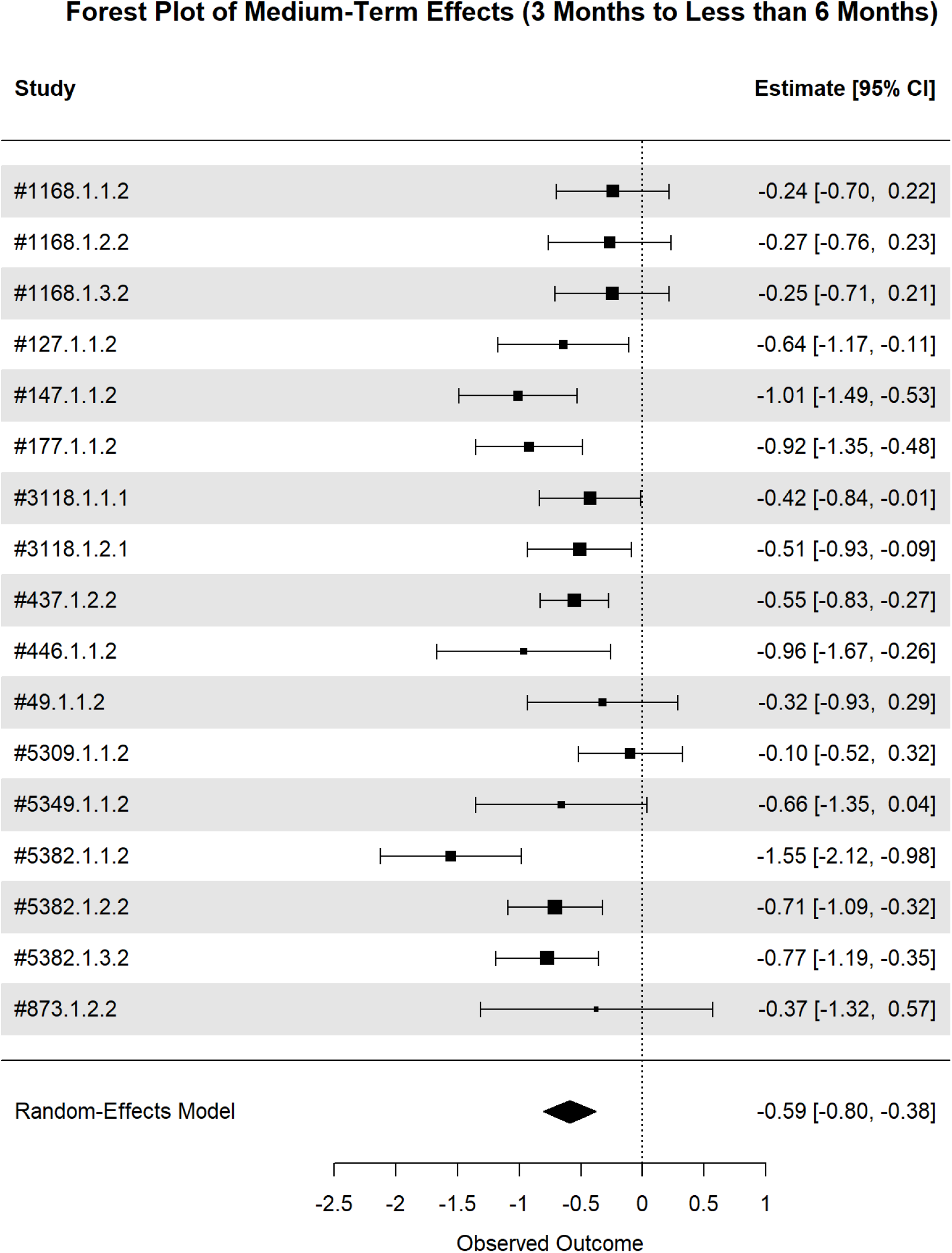
Forest plot for medium-term post-intervention study follow-up timepoints.

For the group of 13 effects measured at or after 6 months post-intervention (see Figure 6), the effect size is small and non-significant (*g* = -0.4370, *p* = 0.0660; 95% CI: -0.9106 to 0.0367). The effects at and after 6 months are highly heterogeneous with an *I*^2^ of 88.2% (*Q =* 81.7, *p* < 0.0001). The tests for publication bias are not statistically significant (Begg and Mazumdar *p =* 0.4354; Egger’s regression intercept *p* = 0.321).

**Figure 6.**
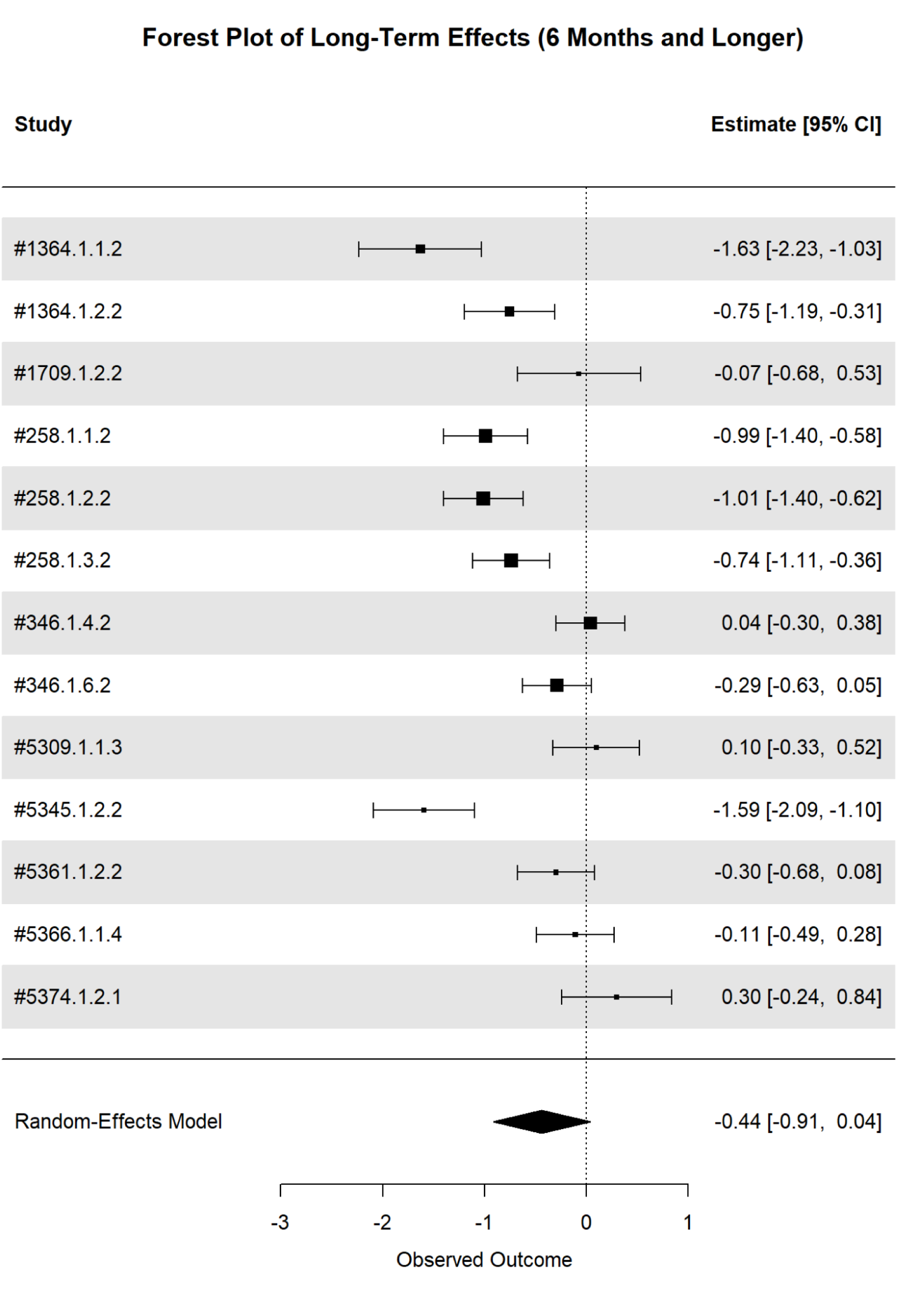
Forest plot for longer-term post-intervention study follow-up timepoints.

### 3.5. Sensitivity analysis results

Removing the nine mid-intervention time points slightly decreases the heterogeneity (*I*^2^ = 78.5%, *Q =* 390.1, *p* < 0.0001). The funnel plot (see Supplemental Figure 2) remains fairly symmetric. Tests for publication bias are non-significant (Begg and Mazumdar *p =* 0.1343; Egger’s regression intercept *p* = 0.5715). There is a small-to-moderate reduction in stress eating with stress management interventions (*g* = -0.4178, *p* < 0.0001; 95% CI: -0.5380 to -0.2977). In a model including study design as a moderator, controlled trials have a small and significant effect (*g =* -0.2102, *p =* 0.0039; 95% CI: -0.3469 to -0.0734). Interventions have a larger effect in pre-post designs, with the effect size increasing in magnitude by 0.4071 (*p* = 0.0003; 95% CI: -0.6179 to -0.1964).

We tested the sensitivity of our results to the definition of “short-term” and “medium-term” effects first by analyzing the 66 effects measured immediately post-intervention separately from other short-term effects. There is a small-to-moderate reduction in stress eating in the immediate measurements (*g* = -0.3997, *p <* 0.0001; 95% CI: -0.5331 to -0.2662). The 14 measurements taken from one week to 2 months post-intervention have a moderate and significant effect (*g* = -0.5855, *p* = 0.0109; 95% CI: -0.9209 to -0.2501). We also assessed effects when considering measurements at 2 months post-intervention to be “medium-term.” In this regrouping, the short-term effect includes 78 measurements from immediate post-intervention to less than 3 months after intervention, and it is small-to-moderate and significant (*g* = -0.4193, *p* < 0.0001; 95% CI: -0.5436 to -0.2950). The medium-term effect, including 19 measurements at 2, 3, and 4 months post-intervention, is moderate and significant (*g* = -0.5886, *p* = 0.0001; 95% CI: -0.8004 to -0.3769).

Removing the 37 effect sizes for which 0.5 is input as the pre-post timepoint correlations reduces the heterogeneity; the *I*^2^ for this analysis is 77.4% (*Q =* 286.4, *p <* 0.0001). The effect size decreases in magnitude slightly but remains significant at -0.3682 (*p <* 0.0001; 95% CI: - 0.5030 to -0.2335). The majority (32 out of 37) of the removed effects are from pre-post designs. Excluding these measurements does not materially impact the analysis with design as a moderator. Controlled trials have a small and significant effect size of -0.1973 (*p* = 0.0088; 95% CI: -0.3401 to -0.0546). The effect increases in magnitude by 0.4573 for pre-post designs (*p =* 0.0004; 95% CI: -0.6903 to -0.2244).

Finally, including the result from one study for which the effect direction was in conflict with the reported pre- and post-treatment means in the published paper in the pooled analysis increases the heterogeneity observed in the pooled analysis; the *I*^2^ is 81.6% (*Q* = 458.4, *p* < 0.0001). The overall effect remains significant with effect size -0.4047 (*p* < 0.0001, 95% CI: - 0.5217 to -0.2876).

## 4. Discussion

Stress eating is a notable obesogenic behavior that engenders obesity risk for women [8,14]. Many existing behavioral weight and lifestyle interventions for obesity frequently target weight loss or management but may fail to explicitly address a formidable challenge to intervention effectiveness due to unaddressed stress eating [13]. Generally, stress eating interventions reduce both stress eating behaviors and body mass index [13–14], and stress management interventions are effective for weight loss [11, 31], yet it is unclear whether stress management interventions reduce stress eating behaviors among women without diagnosed eating disorders. This systematic review and meta-analysis fills this gap in the field’s understanding of the effectiveness of stress management interventions to reduce stress eating for women (Aim 1) and the longevity of any such effects (Aim 2).

Results from this meta-analysis reveal small-to-moderate pooled effects on reducing stress eating, which generally remain consistent in sensitivity analyses. This suggests that interventions that include stress management as an active component are effective in reducing stress eating. Stress management interventions may improve through increasing emotion regulation, reshaping maladaptive cognitions that contribute to stress, and encouraging intentional, nonjudgemental approach of self and situations outside of one’s control [28–30]. The results are generally comparable to previous studies that consider the effects of specific stress management strategies such as mindfulness [14] and stress eating interventions [13], although effect sizes generated in the current meta-analysis were larger than those described or estimated in these previous studies. We intentionally employed a wide inclusion criterion for stress management interventions and the studies included in the review reflect a diversity of intervention delivery methods, components, and durations. Although most included interventions in the current study have acceptance-based stress management components, perhaps the wider inclusion of stress management interventions across the spectrum of approaches indicates a broader effectiveness for stress management strategies writ large. This suggests that it may be a promising future direction to consider widespread inclusion of stress management in existing behavioral and biomedical interventions. Although the effect size was relatively modest, this degree of reduction in stress eating could have meaningful impacts on adherence and success in obesity-related interventions, especially given the prevalence of this challenge for women.

Study design features were examined to make sense of high study heterogeneity. We found stronger effects for pre-post studies without control groups than controlled trials, but the effect of stress management interventions on stress eating were significant for studies of both design types. Aligned with current methodological recommendations for the rigor of controlled trials, it may be harder to demonstrate that stress management interventions create sizable reductions in stress eating using more rigorous designs in which confounds are tightly controlled via randomization and effects must be demonstrated in contrast to other conditions (e.g., waitlist or negative control conditions which may engender reactance or social desirability effects) or forms of treatment (e.g., treatment as usual). Intervention studies included in this review encompassed different types of control groups, which may have contributed to differences in their ability to demonstrate comparative efficacy as a group, thereby reducing the magnitude of the effect size in comparison to pre-post no control group studies. Although the effect size for controlled trials was smaller, stress management interventions still appear to be effective in reducing stress eating in these studies.

Our examination of longevity of effects suggests consistent and significant small-to-moderate effects in the short- and mid-term (i.e., before 6 months) post-intervention. These patterns were generally similar regardless of sensitivity analyses which considered different groupings of follow-up timepoints. Notably, midterm follow-ups (e.g., between 3 and 6 months) demonstrated the largest effect size, suggesting that stress management interventions may have the most profound impact on stress eating in the months following the intervention. One possibility is that, aligned with affect regulation models,[24] stress management interventions are first impacting stress levels, negative affect, or emotion regulation processes more proximally, with the impacts on actual stress eating behavior coming later. Another possibility is that findings may be a function of the distribution of pre-post studies versus controlled trials. In mid-intervention, immediate post-intervention, and long-term follow-up periods, there are relatively more controlled trial effects (∼44%) and effect sizes are smaller. In the short- and medium-term follow-up periods, 36% and 29% of effect sizes came from controlled trials, respectively. These study design differences across these follow-up timepoint groupings could be inflating the effect size estimates, particularly at midterm when the lowest proportion of effects from controlled trials are included.

In line with previous research [13, 42], the effect was no longer significant at the longer follow-up timepoints (6 months or longer), although it was still small in magnitude. This finding extends those of previous reviews which suggest behavioral weight loss interventions (many of which may include stress management components) are effective in the relatively short term [13,42]. Perhaps participants discontinued use of stress management strategies or they became less effective in the longer term. Few studies included in this review measure the degree to which stress management strategies were still utilized or effective for participants at follow-up assessments, so additional research is needed to test this proposition. Our results suggest that stress management interventions for stress eating could be enhanced by testing if booster sessions might be effective in maintenance of reductions in stress eating in the longer term. Design and testing of stress management interventions for stress eating should also incorporate at least a three month follow-up period in order to demonstrate sufficient progress for short-term behavior change, in addition to testing any immediate post-intervention effects. Finally, although fewer studies in our review reported mid-intervention effects, incorporating multiple mid-intervention assessment points when testing and evaluating stress eating interventions is suggested. Such practices could directly inform future research and refinement of studies to determine the intervention dosage that is maximally cost-efficient and effective.

The current study also suggests areas of improvement for equitable inclusion of individuals in research on stress management interventions and stress eating. A key strength of the current investigation is its framing of the importance of these considerations for women’s health particularly and corresponding robust proportion of women in included studies (about 90% on average). Although other studies have examined stress, interventions, and stress eating for women, they have often not set as stringent an inclusion criterion (i.e., at least 70% women), resulting in substantial heterogeneity in the degree to which women are or are not represented in the results. At the same time, our systematic review indicates that existing intervention studies do not frequently include as many older adults or members of minoritized racial/ethnic groups in the United States. Previous research indicates that different age and racial/ethnic groups face different amounts of stress, have different success rates in weight management interventions, and require different intervention and treatment approaches [43]. Therefore, equitable inclusion of these populations in intervention design, testing, and refinement remains a priority to address obesity. Although not an inclusion criterion of ours, studies in this review largely comprised individuals with overweight or obesity. This is appropriate given our focus on stress eating as an important behavioral challenge for women enrolled in weight management interventions, but may limit our understanding of appropriate intervention and prevention strategies for women not engaged in behavioral weight management treatment or interventions for a variety of reasons, both individual (e.g., time commitment, competing roles and responsibilities) and structural (e.g., differential access to treatment, cost). Failure to examine stress eating behaviors among populations without overweight and obesity may also unintentionally serve to further stigmatize and induce internalized weight stigma related to stress eating, although this behavior is not akin to an eating disorder and is engaged by many individuals regardless of health status. Therefore, examination of the effectiveness and accessibility of stress management strategies for stress eating in populations without overweight and obesity remains a priority for future research.

### 4.1. Limitations

Our study is not without limitations. First, although we accounted for high heterogeneity among studies in a variety of ways, a high degree of heterogeneity remained, which could impact the reliability of results. Second, we included tests of risk of bias and heterogeneity during meta-analysis, yet the results largely speak to future intervention research. Clinical practice recommendations should follow a formal GRADE (Grading of Recommendations, Assessment, Development and Evaluation) evaluation. Third, we grouped emotional and stress eating under the general umbrella of stress eating and we considered only emotional eating in response to negative emotions in the current review and meta-analysis. Stress eating and emotional eating, although related, could also be considered distinct in that they tap into different psychosocial propellers of the eating process. Previous research also suggests differences in eating in response to discrete emotions (e.g., sadness versus boredom [44,45]). However, there were not enough studies within each category to examine these differences. We encourage more studies to examine distinct forms of stress eating, which will generate sufficient data to meta-analyze such differences.

Limitations flowing from the features of included studies also constrain the generalizability of results. We explicitly excluded samples composed of only individuals with eating disorders and studies that included only measures of stress eating from eating disorder scales. However, the degree to which samples included individuals without diagnosed eating disorders but with symptoms of eating disorders was not always transparently reported and could not therefore be ascertained. Standardization of reporting of eating disorder symptoms in interventions of eating behaviors is needed to account for underdiagnosis and improve understanding of the degree to which this distinct population requires tailored intervention solutions. Additionally, as noted above, studies in this meta-analysis included most women, individuals with overweight or obesity, White people, and more young and middle adults. Results may not generalize to other groups or an obesity prevention context.

## 5. Conclusion

This meta-analysis found that stress interventions resulted in significant improvements in stress eating. These effects are smaller, yet remain significant, in controlled trials compared to non-equivalent groups pre-test post-test designs. Effects remained significant in the short- and mid- term but were no longer significant at six months and longer post-intervention. Studies of stress eating interventions should strive to incorporate at least a three-month follow-up period and consider ways to boost the effectiveness of stress eating interventions in the longer-term (e.g., Multiphase Optimization Studies (MOST) studies of dosage, intervention components). Examination of study characteristics also suggests new priorities for testing the effectiveness of these interventions among a wider range of individuals. Equitable inclusion of older adults and racial/ethnic minoritized groups in stress management intervention and stress eating studies is suggested. Taken together, results suggest that interventions that target obesity could consider incorporating some aspects of stress management to support women on their health journey.

## Supporting information

Supplemental Materials - Search Strings, PRISMA

## Data Availability

The data that support the findings of this study are available from the corresponding author upon reasonable request.

https://github.com/emdavis7/stress_eating_meta_analysis

## Acknowledgements

None

## Potential conflicts of interest

None

## Footnotes

1 The larger project initially included both correlational and intervention studies. As searching and screening generated sufficient numbers of effect sizes from intervention studies, we only reviewed and meta-analyzed intervention studies in the current investigation. Intervention studies provide the most rigorous evidence base to answer our research questions. Meta-analyses of both intervention and correlational studies would be so heterogeneous they would be unlikely to produce meaningful results.

2 For example, in the long-term group, all pre-post designs are measured at 6 months, while some of the controlled trial effects are measured at a year.

3 During February 2025 when we were conducting our search, the database platform EBSCOhost updated its interface, resulting in significantly more references downloaded into the bibliographic data file than included in the search itself. At the time, the timeline for resolving this known platform issue was uncertain, so we opted to continue with the screening process. This likely resulted in the high number of initial studies screened out as irrelevant, despite our systematic search and screening protocol.

4 One effect, from study #1365, is excluded because the sign of the effect conflicts with reported pre- and post-measurements. This study is included in the sensitivity analysis.

## Notes

### Competing Interest Statement

The authors have declared no competing interest.

