## Supplemental Materials - Search Strings, PRISMA for "Effectiveness of Stress Management to Reduce Stress Eating for Women: A Systematic Review and Meta-analysis of Intervention Studies"

### PRISMA (Preferred Reporting Items for Systematic Reviews and Meta-Analyses) Checklist

www.prisma-statement.org

You must report the page number in your manuscript where you consider each of the items listed in this checklist. If you have not included this information, either revise your manuscript accordingly before submitting or note N/A.

| Section/Topic | Item No. | Checklist item | Reported on Page No. |
|---|---|---|---|
| **TITLE** | | | |
| Title | 1 | Identify the report as a systematic review, meta-analysis, or both. | 1 |
| **ABSTRACT** | | | |
| Structured summary | 2 | Provide a structured summary including, as applicable: background; objectives; data sources; study eligibility criteria, participants, and interventions; study appraisal and synthesis methods; results; limitations; conclusions and implications of key findings; systematic review registration number. | 2 |
| **INTRODUCTION** | | | |
| Rationale | 3 | Describe the rationale for the review in the context of what is already known. | 3 7 |
| Objectives | 4 | Provide an explicit statement of questions being addressed with reference to participants, interventions, comparisons, outcomes, and study design (PICOS). | 7 |
| **METHODS** | | | |
| Protocol and registration | 5 | Indicate if a review protocol exists, if and where it can be accessed (e.g., Web address), and, if available, provide registration information including registration number. | 7 |
| Eligibility criteria | 6 | Specify study characteristics (e.g., PICOS, length of follow-up) and report characteristics (e.g., years considered, language, publication status) used as criteria for eligibility, giving rationale. | 8 9 |
| Information sources | 7 | Describe all information sources (e.g., databases with dates of coverage, contact with study authors to identify additional studies) in the search and date last searched. | 9 10, |
| Search | 8 | Present full electronic search strategy for at least one database, including any limits used, such that it could be repeated. | Supplement |
| Study selection | 9 | State the process for selecting studies (i.e., screening, eligibility, included in systematic review, and, if applicable, included in the meta-analysis). | 10 12 |
| Data collection process | 10 | Describe method of data extraction from reports (e.g., piloted forms, independently, in duplicate) and any processes for obtaining and confirming data from investigators. | 10 11 |
| Data items | 11 | List and define all variables for which data were sought (e.g., PICOS, funding sources) and any assumptions and simplifications made. | 9,11,12 |
| Risk of bias in individual studies | 12 | Describe methods used for assessing risk of bias of individual studies (including specification of whether this was done at the study or outcome level), and how this information is to be used in any data synthesis. | 14 |

| Section/Topic | Item No. | Checklist item | Reported on Page No. |
|---|---|---|---|
| Summary measures | 13 | State the principal summary measures (e.g., risk ratio, difference in means). | 12 |
| Synthesis of results | 14 | Describe the methods of handling data and combining results of studies, if done, including measures of consistency (e.g., $I^2$) for each meta-analysis. | 11 13 |
| Risk of bias across studies | 15 | Specify any assessment of risk of bias that may affect the cumulative evidence (e.g., publication bias, selective reporting within studies). | 14 |
| Additional analyses | 16 | Describe methods of additional analyses (e.g., sensitivity or subgroup analyses, meta-regression), if done, indicating which were pre-specified. | 13 14 |
| **RESULTS** | | | |
| Study selection | 17 | Give numbers of studies screened, assessed for eligibility, and included in the review, with reasons for exclusions at each stage, ideally with a flow diagram. | 14 15, 35 |
| Study characteristics | 18 | For each study, present characteristics for which data were extracted (e.g., study size, PICOS, follow-up period) and provide the citations. | 15 18, Supp |
| Risk of bias within studies | 19 | Present data on risk of bias of each study and, if available, any outcome level assessment (see item 12). | N/A |
| Results of individual studies | 20 | For all outcomes considered (benefits or harms), present, for each study: (a) simple summary data for each intervention group (b) effect estimates and confidence intervals, ideally with a forest plot. | 37 40 |
| Synthesis of results | 21 | Present results of each meta-analysis done, including confidence intervals and measures of consistency. | 18 20 |
| Risk of bias across studies | 22 | Present results of any assessment of risk of bias across studies (see Item 15). | 18 20,36 |
| Additional analysis | 23 | Give results of additional analyses, if done (e.g., sensitivity or subgroup analyses, meta-regression [see Item 16]). | 20 21 |
| **DISCUSSION** | | | |
| Summary of evidence | 24 | Summarize the main findings including the strength of evidence for each main outcome; consider their relevance to key groups (e.g., healthcare providers, users, and policy makers). | 21 26 |
| Limitations | 25 | Discuss limitations at study and outcome level (e.g., risk of bias), and at review-level (e.g., incomplete retrieval of identified research, reporting bias). | 26 27 |
| Conclusions | 26 | Provide a general interpretation of the results in the context of other evidence, and implications for future research. | 27 |
| **FUNDING** | | | |

| Section/Topic | Item No. | Checklist item | Reported on Page No. |
|---|---|---|---|
| Funding | 27 | Describe sources of funding for the systematic review and other support (e.g., supply of data); role of funders for the systematic review. | 1 |

*From:* Moher D, Liberati A, Tetzlaff J, Altman DG, The PRISMA Group (2009). Preferred Reporting Items for Systematic Reviews and Meta-Analyses: The PRISMA Statement. PLoS Med 6(6): e1000097. doi:10.1371/journal.pmed1000097

**Once you have completed this checklist, please save a copy and upload it as part of your submission. Please DO NOT include this checklist as part of the main manuscript document. It must be uploaded as a separate file.**

**Supplemental Material 2**

**EBSCOhost (indexes APA PsycInfo, CINAHL):**

(“Stress management” OR “stress reduction” OR “coping with stress” OR (stress AND psychoeducation) OR (stress AND “skill* building”) OR (stress AND “skill* acquisition”) OR (stress and “cognitive reappraisal”) OR mindfulness OR compassion OR relaxation OR biofeedback OR acceptance* OR “emotion regulation” OR (stress AND “cognitive therapy”) OR (stress AND “cognitive reframing”) (stress AND “cognitive restructuring”) OR (stress AND problem-solving) OR (stress AND “Cognitive behavi#ral therapy”) OR (stress AND “Dialectical behavi#r therapy”) OR (stress AND “Acceptance and commitment therapy”) OR (stress AND inoculation) OR (stress AND “attention and interpretation therapy”) OR (stress and “coping skills”) OR “stress-related”) **AND** (“Stress eating” OR “stress-induced eating” OR “stress-related eating” OR “stress eating practice*” OR “emotional eating” OR “eating to cope” OR “eating in response to negative emotions” OR “coping using eating” OR “emotional overeating”) **AND** (Women* OR “Woman*” OR “male-to-female*” OR “femme*” OR “Female*” OR “Adult*Women*”)

**PubMed:**

“Stress management” OR “stress reduction” OR “coping with stress” OR (stress AND psychoeducation) OR (stress and “skill* building”) OR (stress and “skill* acquisition”) OR (stress and “cognitive reappraisal”) OR mindfulness OR compassion OR relaxation OR biofeedback OR acceptance* OR “emotion regulation” OR (stress AND “cognitive therapy”) OR (stress AND “cognitive reframing”) (stress AND “cognitive restructuring”) OR (stress AND problem-solving) OR (stress AND “Cognitive behavioral therapy”) OR (stress AND “Dialectical behavior therapy”) OR (stress AND “Acceptance and commitment therapy”) OR (stress AND inoculation) OR (stress AND “attention and interpretation therapy”) OR (stress and “coping skills”) OR “stress-related”) **AND** (“Stress eating” OR “stress-induced eating” OR “stress-related eating” OR “stress eating practice*” OR “emotional eating” OR “eating to cope” OR “eating in response to negative emotions” OR “coping using eating” OR “emotional overeating”) **AND** (Women* OR “Woman*” OR “male-to-female*” OR “femme*” OR “Female*” OR “Adult*Women*”)

MeSH

- mindfulness"[MeSH Terms]
- "empathy"[MeSH Terms]
- relaxation"[MeSH Terms]
- "biofeedback, psychology"[MeSH Terms]
- problem solving"[MeSH Terms]
- vaccination"[MeSH Terms]
- attention"[MeSH Terms]
- therapeutics"[MeSH Terms]
- therapy"[MeSH Subheading]

**Web of Science (indexes Medline):**

("Stress management" OR "stress reduction" OR "coping with stress") OR (stress AND psychoeducation) OR (stress and "skill* building") OR (stress and "skill* acquisition") OR (stress and "cognitive reappraisal") OR (mindfulness OR compassion OR relaxation OR biofeedback OR acceptance* OR "emotion regulation") OR (stress AND "cognitive therapy") OR (stress AND "cognitive reframing") OR (stress AND "cognitive restructuring") OR (stress AND problem-solving) OR (stress AND "Cognitive behavi$ral therapy") OR (stress AND "Dialectical behavi$r therapy") OR (stress AND "Acceptance and commitment therapy") OR (stress AND inoculation) OR (stress AND "attention and interpretation therapy") OR (stress and "coping skills") OR ("stress-related") **AND** "Stress eating" OR "stress-induced eating" OR "stress-related eating" OR "stress eating practice*" OR "emotional eating" OR "eating to cope" OR "eating in response to negative emotions" OR "coping using eating" OR "emotional overeating") **AND** (Women* OR "Woman*" OR "male-to-female*" OR "femme*" OR "Female*" OR "Adult*Women*")

### Supplemental Material 3

In this section, we give additional details on the multi-level meta-analysis. Within a particular study $i$ ($i$ = 1,…, 60), we may record outcomes from multiple stress management interventions ($j$; $j$ = 1,…, $n_i$). Each intervention may track more than one outcome, $k$ ($k$ = 1,…, $n_{ij}$). Lastly, an outcome may report effects at multiple time points post-intervention, $l$ ($l$ = 1,..., $n_{ijk}$).

As in a standard random-effects meta-analysis, our multi-level meta-analysis estimates $\mu$ in the following equation

$$Y = \mu \, 1_N + \varepsilon,$$

where $\boldsymbol{Y}$ is the vector of observed effect sizes, $\mu$ is a scalar pooled effect of interest, $\mathbf{1}_N$ is a vector of ones, and $\varepsilon \sim N(\mathbf{0}, \mathbf{\Sigma})$. In our analysis, the covariance matrix $\mathbf{\Sigma}$ is block-diagonal with the following covariance terms:

The variance of each observation is

$$Cov(Y_{ijkl}, Y_{ijkl}) = \sigma^2{}_{study} + \sigma^2{}_{int} + \sigma^2{}_{outcome} + \sigma^2{}_{time} + \sigma^2{}_{residual}$$

For observations with the same study, intervention, and outcome, but at different timepoints,

$$Cov(Y_{ijkl}, Y_{ijkl'}) = \sigma^2{}_{study} + \sigma^2{}_{int} + \sigma^2{}_{outcome} + \sigma^2{}_{time}\rho^{|l - l'|}$$

For observations from the same study and intervention, but measuring different outcomes,

$$Cov(Y_{ijkl}, Y_{ijk'l}) = \sigma^2{}_{study} + \sigma^2{}_{int}$$

For observations from the same study, but with different interventions,

$$Cov(Y_{ijkl}, Y_{ij'kl}) = \sigma^2{}_{study}$$

Lastly, observations from different studies are assumed to be independent:

$$Cov(Y_{i'jkl}, Y_{ijkl}) = 0$$

We estimate these covariance parameters and $\mu$ using restricted maximum likelihood, with the metafor package in R.

### Supplemental Material 4

#### List of References for Included Studies

Afari N, Herbert MS, Godfrey KM, et al. Acceptance and commitment therapy as an adjunct to the MOVE! programme: a randomized controlled trial. Obesity Science & Practice. 2019;5(5):397-407. doi:10.1002/osp4.356 **(Study #5309)**

Alberts HJEM, Mulkens S, Smeets M, Thewissen R. Coping with food cravings. Investigating the potential of a mindfulness-based intervention. Appetite. 2010;55(1):160-163. doi:10.1016/j.appet.2010.05.044 **(Study #5299)**

Alberts HJEM, Thewissen R, Raes L. Dealing with problematic eating behaviour. The effects of a mindfulness-based intervention on eating behaviour, food cravings, dichotomous thinking and body image concern. Appetite. 2012;58(3):847-851. doi:10.1016/j.appet.2012.01.009 **(Study #429)**

Anand C, Hengst K, Gellner R, Englert H. Effects of the healthy lifestyle community program (cohort 1) on stress-eating and weight change after 8 weeks: a controlled study. Sci Reports. 2023;13(1). doi:10.1038/s41598-022-27063-4 **(Study #1367)**

Annesi JJ. Effects of cardiovascular exercise on eating behaviours: Accounting for effects on stress, depression-, and anger-related emotional eating in women with obesity. Stress Health. 2024;40(4):e3364. doi:10.1002/smi.3364 **(Study #898)**

Annesi JJ. Relationship of Emotional Eating and Mood Changes Through Self-Regulation Within Three Behavioral Treatments for Obesity. Psychol Rep. 2019;122(5):1689-1706. doi:10.1177/0033294118795883 **(Study #5366)**

Annesi JJ, Mareno N, McEwen K. Psychosocial predictors of emotional eating and their weight-loss treatment-induced changes in women with obesity. Eat Weight Disord. 2016;21(2):289-295. doi:10.1007/s40519-015-0209-9 **(Study #5381)**

Beaulac J, Sandre D, Mercer D. Impact on mindfulness, emotion regulation, and emotional overeating of a DBT skills training group: a pilot study. Eat Weight Disord. 2019;24(2):373-377. doi:10.1007/s40519-018-0616-9 **(Study #446)**

Boucher S, Edwards O, Gray A, et al. Teaching Intuitive Eating and Acceptance and Commitment Therapy Skills Via a Web-Based Intervention: A Pilot Single-Arm Intervention Study. JMIR Research Protocols. 2016;5(4):e5861. doi:10.2196/resprot.5861 **(Study #5351)**

Braden A, O'Brien W. Pilot Study of a Treatment Using Dialectical Behavioral Therapy Skills for Adults with Overweight/Obesity and Emotional Eating. J Contemp Psychother. 2021;51(1):21-29. doi:10.1007/s10879-020-09477-1 **(Study #5370)**

Braden A, Redondo R, Ferrell E, et al. An Open Trial Examining Dialectical Behavior Therapy Skills and Behavioral Weight Loss for Adults With Emotional Eating and Overweight/Obesity. Behav Therapy. 2022;53(4):614-627. doi:10.1016/j.beth.2022.01.008 **(Study #258)**

Bradley LE, Forman EM, Kerrigan SG, et al. Project HELP: a Remotely Delivered Behavioral Intervention for Weight Regain after Bariatric Surgery. Obes Surg. 2017;27(3):586-598. doi:10.1007/s11695-016-2337-3 **(Study #5421)**

Campos M, Menezes I, Peixoto M, Schincaglia R. Intuitive eating in general aspects of eating behaviors in individuals with obesity: Randomized clinical trial. Clin Nutr NESP. 2022;50:24-32. doi:10.1016/j.clnesp.2022.06.002 **(Study #1709)**

Cancian ACM, de Souza LAS, Liboni RPA, Machado W de L, Oliveira M da S. Effects of a dialectical behavior therapy-based skills group intervention for obese individuals: a Brazilian pilot study. Eat Weight Disord. 2019;24(6):1099-1111. doi:10.1007/s40519-017-0461-2 **(Study #186)**

Chacko SA, Yeh GY, Davis RB, Wee CC. A mindfulness-based intervention to control weight after bariatric surgery: Preliminary results from a randomized controlled pilot trial. Complement Thera Med. 2016;28:13-21. doi:10.1016/j.ctim.2016.07.001 **(Study #49)**

Christaki E, Kokkinos A, Costarelli V, Alexopoulos EC, Chrousos GP, Darviri C. Stress management can facilitate weight loss in Greek overweight and obese women: a pilot study. J Human Nutr Diet. 2013;26 Suppl 1:132-139. doi:10.1111/jhn.12086 **(Study #1292)**

Czepczor-Bernat K, Brytek-Matera A, Staniszewska A. The effect of a web-based psychoeducation on emotional functioning, eating behaviors, and body image among premenopausal women with excess body weight. Arch Women Mental Health. 2021;24(3):423-435. doi:10.1007/s00737-020-01077-1 **(Study #120)**

Di Sante J, Frayn M, Angelescu A, Knauper B. Proof-of-concept testing of a brief virtual ACT workshop for emotional eating. Appetite. 2024;199:107386-107386. doi:10.1016/j.appet.2024.107386 **(Study #147)**

Epel E, Laraia B, Coleman-Phox K, et al. Effects of a Mindfulness-Based Intervention on Distress, Weight Gain, and Glucose Control for Pregnant Low-Income Women: A Quasi-Experimental Trial Using the ORBIT Model. Intl J Behav Med. 2019;26(5):461-473. doi:10.1007/s12529-019-09779-2 **(Study #146)**

Felske AN, Williamson TM, Rash JA, Telfer JA, Toivonen KI, Campbell T. Proof of Concept for a Mindfulness-Informed Intervention for Eating Disorder Symptoms, Self-Efficacy, and Emotion Regulation among Bariatric Surgery Candidates. Behav Med. 2022;48(3):216-229. doi:10.1080/08964289.2020.1828255 **(Study #437)**

Forman E, Butryn M, Hoffman K, Herbert J. An Open Trial of an Acceptance-Based Behavioral Intervention for Weight Loss. Cog Behav Pract. 2009;16(2):223-235. doi:10.1016/j.cbpra.2008.09.005 **(Study #1582)**

Frayn M, Khanyari S, Knäuper B. A 1-day acceptance and commitment therapy workshop leads to reductions in emotional eating in adults. Eat Weight Disord. 2020;25(5):1399-1411. doi:10.1007/s40519-019-00778-6 **(Study #5382)**

Haley E, Dolbier C, Carels R, Whited M. A brief pilot self-compassion intervention for women with overweight/obesity and internalized weight bias: Feasibility, acceptability, and future directions. J Context Behav Sci. 2022;23:59-63. doi:10.1016/j.jcbs.2021.12.001 **(Study #1365)**

Haley EN, Dolbier CL, Campbell LC, Carels RA, Braciszewski JM. Brief Self-Compassion Intervention for Women of Higher Weight and Internalized Weight Bias: A Randomized Pilot Study. Intl J Behav Med. Published online 2024. doi:10.1007/s12529-024-10297-z **(Study #716)**

Jarvela-Reijonen E, Karhunen L, Sairanen E, et al. The effects of acceptance and commitment therapy on eating behavior and diet delivered through face-to-face contact and a mobile app: a randomized controlled trial. Intl J Behav Nutr Phys Act. 2018;15(1):22-22. doi:10.1186/s12966-018-0654-8 **(Study #346)**

Jastreboff AM, Chaplin TM, Finnie S, et al. Preventing Childhood Obesity Through a Mindfulness-Based Parent Stress Intervention: A Randomized Pilot Study. J Pediatrics. 2018;202:136-142.e1. doi:10.1016/j.jpeds.2018.07.011 **(Study #59)**

Jiying Ling, Sisi Chen, Nanhua Zhang, Robbins LB, Kerver JM. Happy Family, Healthy Kids: A Healthy Eating and Stress Management Program in Low-Income Parent-Preschooler Dyads. Nurs Res. 2024;73(1):3-15. doi:10.1097/NNR.0000000000000697 **(Study #2292)**

Juarascio AS, Parker MN, Manasse SM, Barney JL, Wyckoff EP, Dochat C. An exploratory component analysis of emotion regulation strategies for improving emotion regulation and emotional eating. Appetite. 2020;150:104634-104634. doi:10.1016/j.appet.2020.104634 **(Study #370)**

Katterman SN, Goldstein SP, Butryn ML, Forman EM, Lowe MR. Efficacy of an acceptance-based behavioral intervention for weight gain prevention in young adult women. J Context Behav Sci. 2014;3(1):45-50. doi:10.1016/j.jcbs.2013.10.003 **(Study #5350)**

Knol L, Appel S, Crowe-White K, Brantley C, Adewumi O, Senkus K. Development, Feasibility, and Initial Results of a Mindful Eating Intervention: Project Mindful Eating and Exercise (MEE): Feeding the Mind, Body, and Soul. Am J Health Edu. 2021;52(4):171-184. doi:10.1080/19325037.2021.1930615 **(Study #1080)**

Laird B, Puzia M, Larkey L, Ehlers D, Huberty J. A Mobile App for Stress Management in Middle-Aged Men and Women (Calm): Feasibility Randomized Controlled Trial. JMIR Form Res. 2022;6(5):e30294. doi:10.2196/30294 **(Study #803)**

Laraia BA, Adler NE, Coleman-Phox K, et al. Novel Interventions to Reduce Stress and Overeating in Overweight Pregnant Women: A Feasibility Study. Mat Child Health J. 2018;22(5):670-678. doi:10.1007/s10995-018-2435-z **(Study #390)**

Lattimore P. Mindfulness-based emotional eating awareness training: taking the emotional out of eating. Eat Weight Disord. 2020;25(3):649-657. doi:10.1007/s40519-019-00667-y **(Study #415)**

Ledoux T, Gallagher MR, Ciampolini M, Sampson M. Biofeedback enhanced lifestyle intervention: exploring the experience of participants in a novel intervention for disinhibited eating and obesity. Open J Prev Med. 2014;4(10):779-788. **(Study #3141)**

Levin ME, Potts S, Haeger J, Lillis J. Delivering Acceptance and Commitment Therapy for Weight Self-Stigma Through Guided Self-Help: Results From an Open Pilot Trial. Cog Behav Practice. 2018;25(1):87-104. doi:10.1016/j.cbpra.2017.02.002 **(Study #5349)**

Marques CC, Palmeira L, Castilho P, et al. Online Compassion Focused Therapy for overeating: Feasibility and acceptability pilot study. Intl J Eat Disord. 2024;57(2):410-422. doi:10.1002/eat.24118 **(Study #127)**

Mason C, de Dieu Tapsoba J, Duggan C, Wang CY, Alfano CM, McTiernan A. Eating behaviors and weight loss outcomes in a 12-month randomized trial of diet and/or exercise intervention in postmenopausal women. Int J Behav Nutr Phys Act. 2019;16(1):113. doi:10.1186/s12966-019-0887-1 **(Study #5363)**

Medina J, Hopkins L, Powers M, Baird S, Smits J. The Effects of a Hatha Yoga Intervention on Facets of Distress Tolerance. Cog Behav Therapy. 2015;44(4):288-300. doi:10.1080/16506073.2015.1028433 **(Study #1489)**

Mensinger J, Shepherd B, Schapiro S, et al. Mediating effects of a weight-inclusive health promotion program on maladaptive eating in women with high body mass index. Eat Behav. 2023;49. doi:10.1016/j.eatbeh.2023.101730 **(Study #1152)**

Mohseni M, Kuckuck S, Meeusen REH, et al. Improved Physical and Mental Health After a Combined Lifestyle Intervention with Cognitive Behavioural Therapy for Obesity. Int J Endocrinol Metab. 2022;21(1):e129906. doi:10.5812/ijem-129906 **(Study #5415)**

Moraes A dos S, Padovani R da C, La Scala Teixeira CV, et al. Cognitive Behavioral Approach to Treat Obesity: A Randomized Clinical Trial. Front Nutr. 2021;8. doi:10.3389/fnut.2021.611217 **(Study #5374)**

Moreira MFS, de Azevedo BEF, Beretta MV, Busnello FM. Nutritional Counseling Based on Mindful Eating for the Eating Behavior of People Living with Overweight and Obesity: A Randomized Clinical Trial. Nutrients. 2024;16(24). doi:10.3390/nu16244388 **(Study #361)**

Paans NPG, Bot M, Brouwer IA, et al. Effects of food-related behavioral activation therapy on eating styles, diet quality and body weight change: Results from the MooDFOOD Randomized Clinical Trial. J Psychosom Res. 2020;137:110206. doi:10.1016/j.jpsychores.2020.110206 **(Study #5372)**

Palazzo C, Leghi B, Diez-Garcia R. Does feeling what you eat change how you eat? Implications of an intervention to promote consciousness of eating experiences. Frontiers Psychol. 2024;14. doi:10.3389/fpsyg.2023.1229105 **(Study #1700)**

Palmeira L, Cunha M, Pinto-Gouveia J. Processes of change in quality of life, weight self-stigma, body mass index and emotional eating after an acceptance-, mindfulness- and compassion-based group intervention (Kg-Free) for women with overweight and obesity. J Health Psychol. 2019;24(8):1056-1069. doi:10.1177/1359105316686668 **(Study #177)**

Palmeira L, Pinto-Gouveia J, Cunha M. Exploring the efficacy of an acceptance, mindfulness & compassionate-based group intervention for women struggling with their weight (Kg-Free): A randomized controlled trial. Appetite. 2017;112:107-116. doi:10.1016/j.appet.2017.01.027 **(Study #5346)**

Paul L, van der Heiden C, van Hoeken D, et al. Cognitive Behavioral Therapy Versus Usual Care Before Bariatric Surgery: One-Year Follow-Up Results of a Randomized Controlled Trial. Obes Surg. 2021;31(3):970-979. doi:10.1007/s11695-020-05081-3 **(Study #5361)**

Pérez C, Cruzat-Mandich C, Bergeret A, et al. Comparative efficacy of remotely delivered mindfulness-based eating awareness training versus behavioral-weight loss counseling during COVID-19. Frontiers Psychol. 2023;14. doi:10.3389/fpsyg.2023.1101120 **(Study #1068)**

Pimenta F, Leal I, Maroco J, Ramos C. Brief cognitive-behavioral therapy for weight loss in midlife women: a controlled study with follow-up. Int J Womens Health. 2012;4:559-567. doi:10.2147/IJWH.S35246 **(Study #873)**

Potts S, Krafft J, Levin M. A Pilot Randomized Controlled Trial of Acceptance and Commitment Therapy Guided Self-Help for Overweight and Obese Adults High in Weight Self-Stigma. Behav Mod. 2022;46(1):178-201. doi:10.1177/0145445520975112 **(Study #1546)**

Roosen MA, Safer D, Adler S, Cebolla A, van Strien T. Group dialectical behavior therapy adapted for obese emotional eaters; a pilot study. Nutricion Hospitalaria. 2012;27(4):1141-1147. **(Study #5345)**

Rychescki GG, Dos Santos GR, Bertin CF, et al. Online Cognitive-Behavioral Therapy-Based Nutritional Intervention via Instagram for Overweight and Obesity. Nutrients. 2024;16(23). doi:10.3390/nu16234045 **(Study #788)**

Sockalingam S, Leung SE, Hawa R, et al. Telephone-based cognitive behavioural therapy for female patients 1-year post-bariatric surgery: A pilot study. Obes Res Clinic Practice. 2019;13(5):499-504. doi:10.1016/j.orcp.2019.07.003 **(Study #2830)**

Soriano-Ayala E, Amutio A, Franco C, Manas I. Promoting a Healthy Lifestyle through Mindfulness in University Students: A Randomized Controlled Trial. Nutrients. 2020;12(8). doi:10.3390/nu12082450 **(Study #27)**

Southgate D, Greiver M, Hubka G, et al. Effect of a Group Behavioural Management Program on Emotional Regulation of Food Choices: A Pilot Randomized Controlled Trial. Canad J Diet Practice Res. 2017;78(3):137-140. doi:10.3148/cjdpr-2017-006 **(Study #655)**

Tapper K, Shaw C, Ilsley J, Hill AJ, Bond FW, Moore L. Exploratory randomised controlled trial of a mindfulness-based weight loss intervention for women. Appetite. 2009;52(2):396-404. **(Study #3118)**

Thomas EA, Mijangos JL, Hansen PA, et al. Mindfulness-Oriented Recovery Enhancement Restructures Reward Processing and Promotes Interoceptive Awareness in Overweight Cancer Survivors: Mechanistic Results From a Stage 1 Randomized Controlled Trial. Integrat Cancer Therapies. 2019;18:1534735419855138-1534735419855138. doi:10.1177/1534735419855138 **(Study #682)**

Timmerman GM, Brown A. The Effect of a Mindful Restaurant Eating Intervention on Weight Management in Women. J Nutr Educ Behav. 2012;44(1):22-28. doi:10.1016/j.jneb.2011.03.143 **(Study #5306)**

Tuncer G, Duman Z. Effects of online-guided group self-help program on female nursing students' attempts to cope with their emotional eating and uncontrolled eating behaviors: A quasi-experimental study. J Psychiat Nurs. 2024;15(2):120-131. doi:10.14744/phd.2023.54926 **(Study #1364)**

Vela AM, Palmer B, Gil-Rivas V, Cachelin F. The Role of Disordered Eating in Type 2 Diabetes: A Pilot Study. Am J Lifestyle Med. 2021;17(1):131-139. doi:10.1177/15598276211002459 **(Study #3521)**

Wang W, Ding X. A pilot randomized trial of self-compassion writing for young adult women engaged in emotional eating in the context of appearance-related cyberbullying. Intl J Eat Dis. 2023;56(8):1520-1533. doi:10.1002/eat.23967 **(Study #139)**

Ward K, Herekar A, Wang P, Lindsay KL. Feasibility and Acceptability of a Mindfulness-Based Smartphone App among Pregnant Women with Obesity. Int J Environ Res Pub Health. 2023;20(7). doi:10.3390/ijerph20075421 **(Study #279)**

Wnuk S, Du C, Van Exan J, et al. Mindfulness-Based Eating and Awareness Training for Post-Bariatric Surgery Patients: a Feasibility Pilot Study. Mindfulness. 2018;9(3):949-960. doi:10.1007/s12671-017-0834-7 **(Study #1168)**

### Supplemental Figures

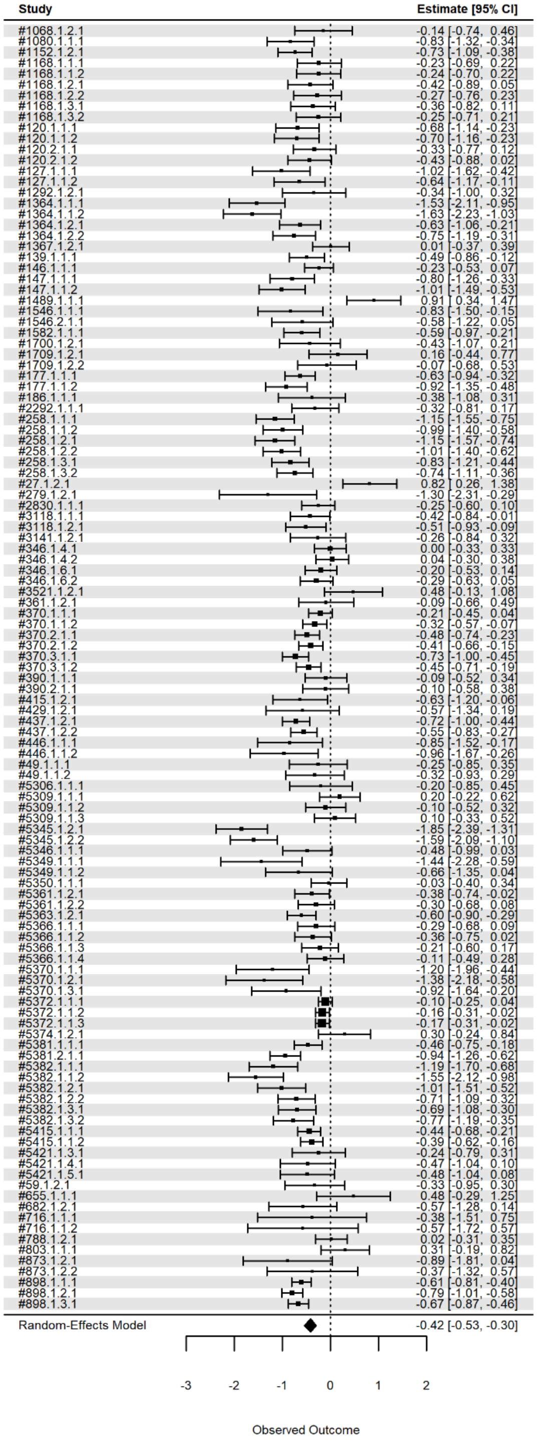


**Supplemental Figure 1**. Forest plot for all studies.

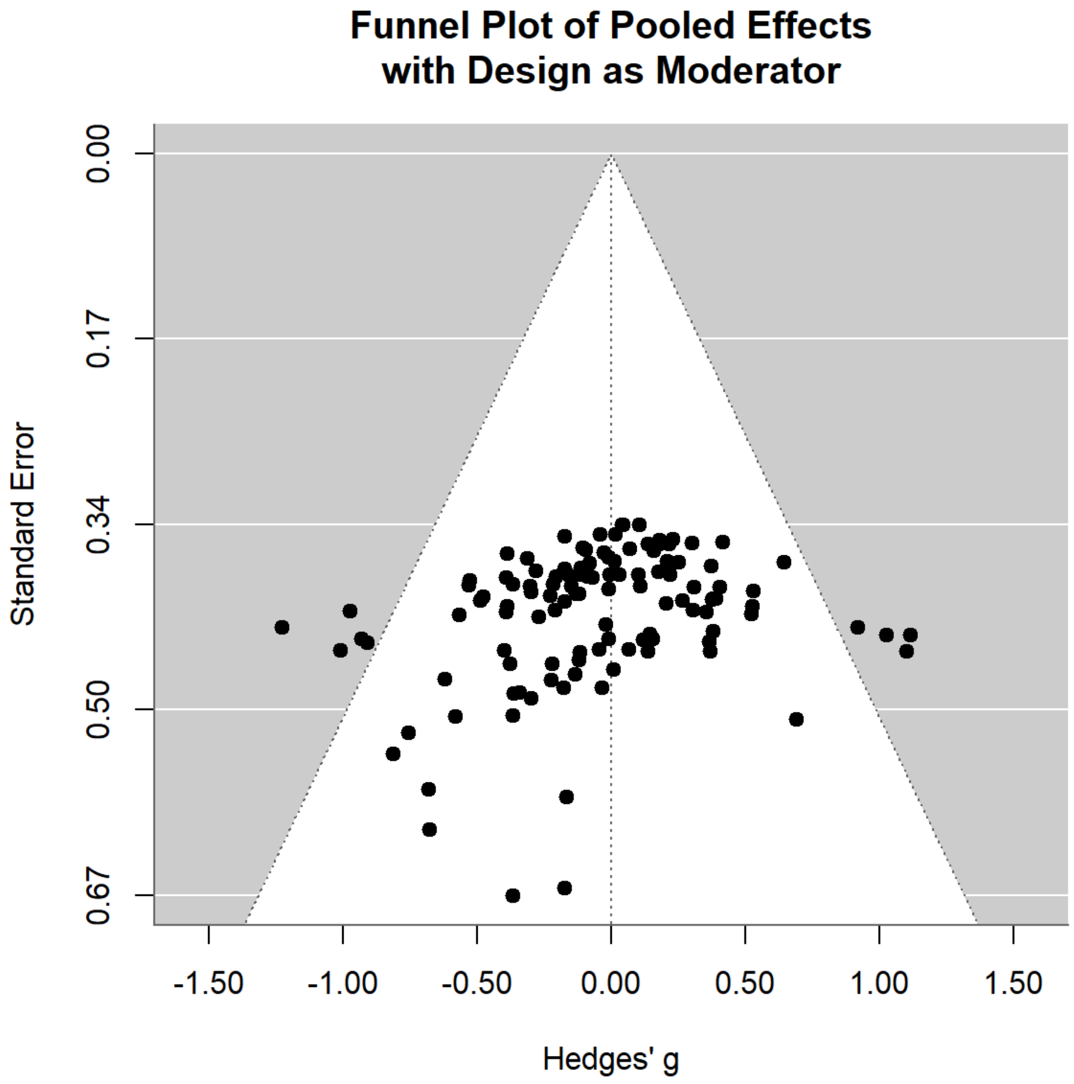


**Supplemental Figure 2.** The funnel plot for pooled effects, when using the design of the study as a moderator.

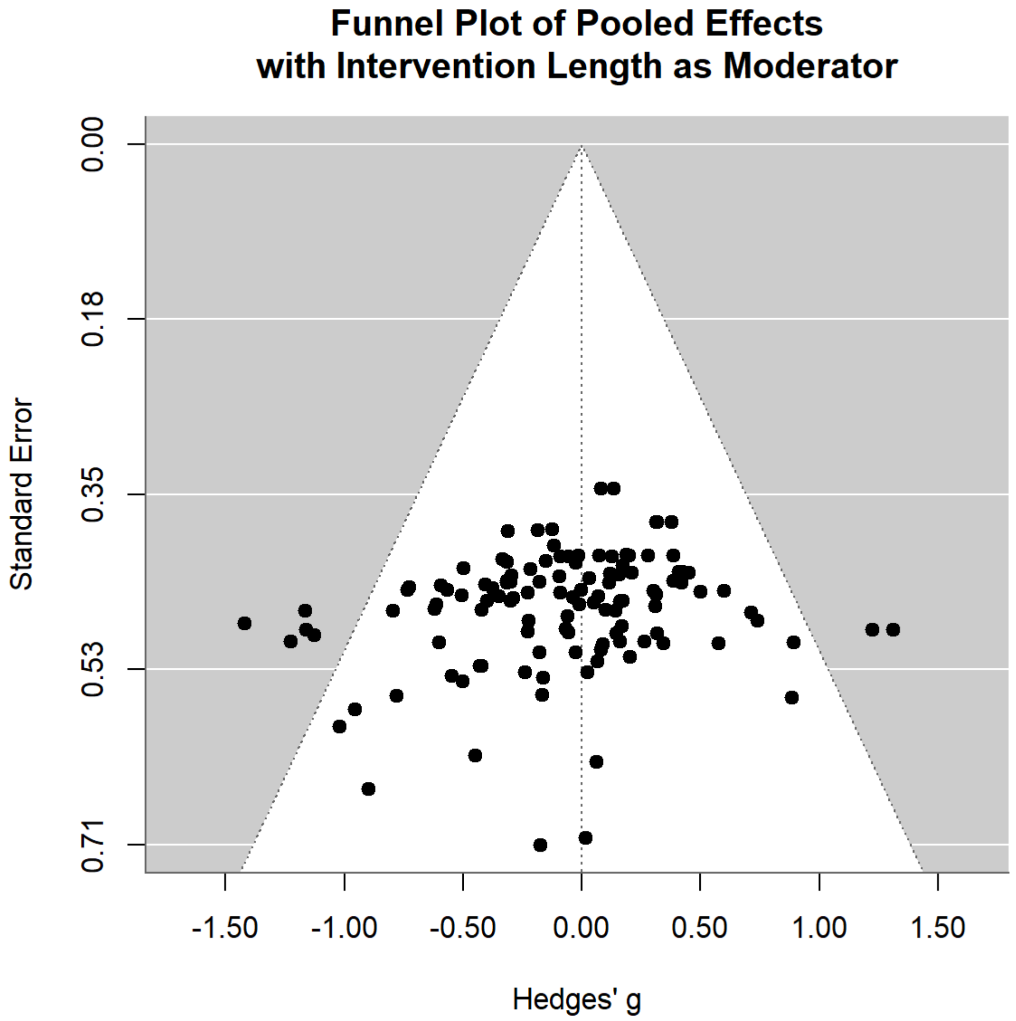


**Supplemental Figure 3.** The funnel plot for pooled effects, when using the intervention length as a moderator.

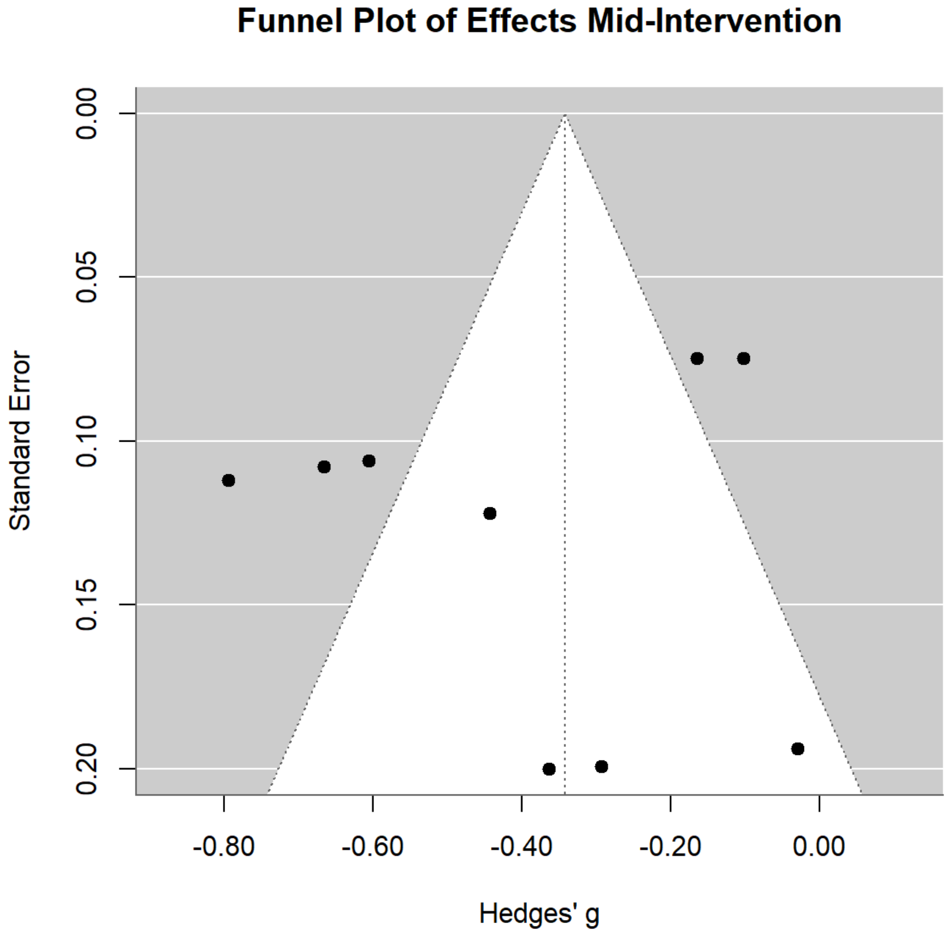


**Supplemental Figure 4.** The funnel plot for effects measured mid-intervention.

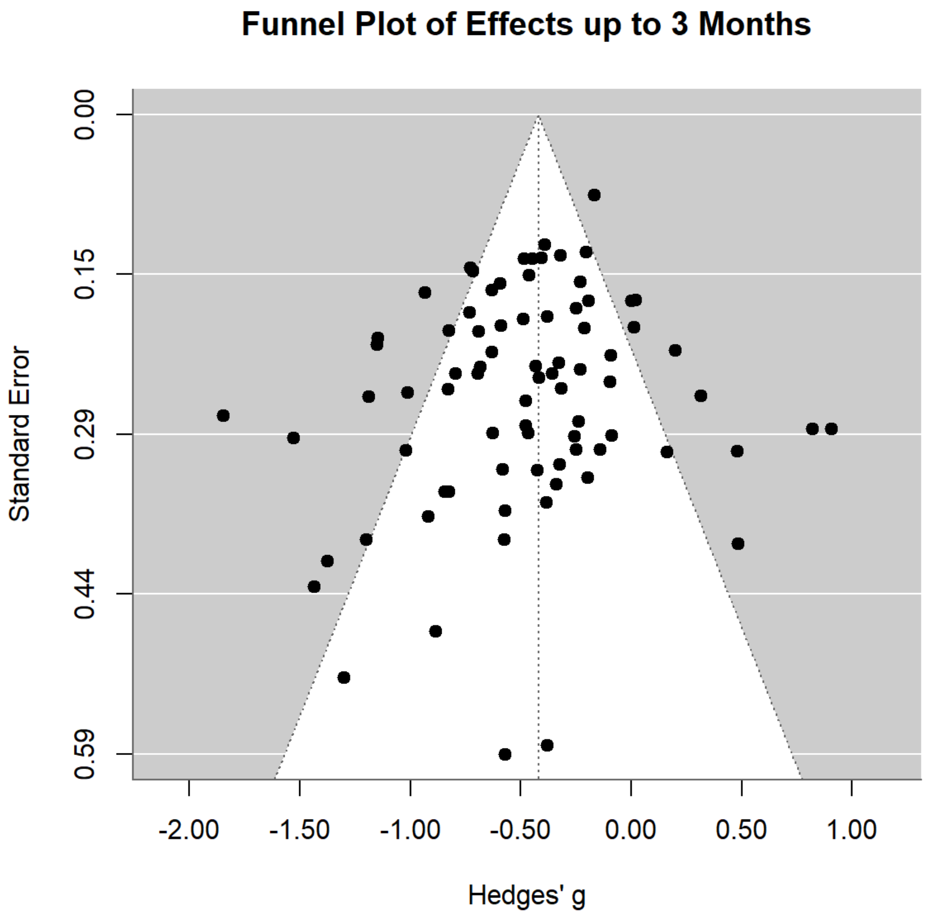


**Supplemental Figure 5.** The funnel plot for effects measured in the short-term (from immediately post-intervention to less than 3 months post-intervention).

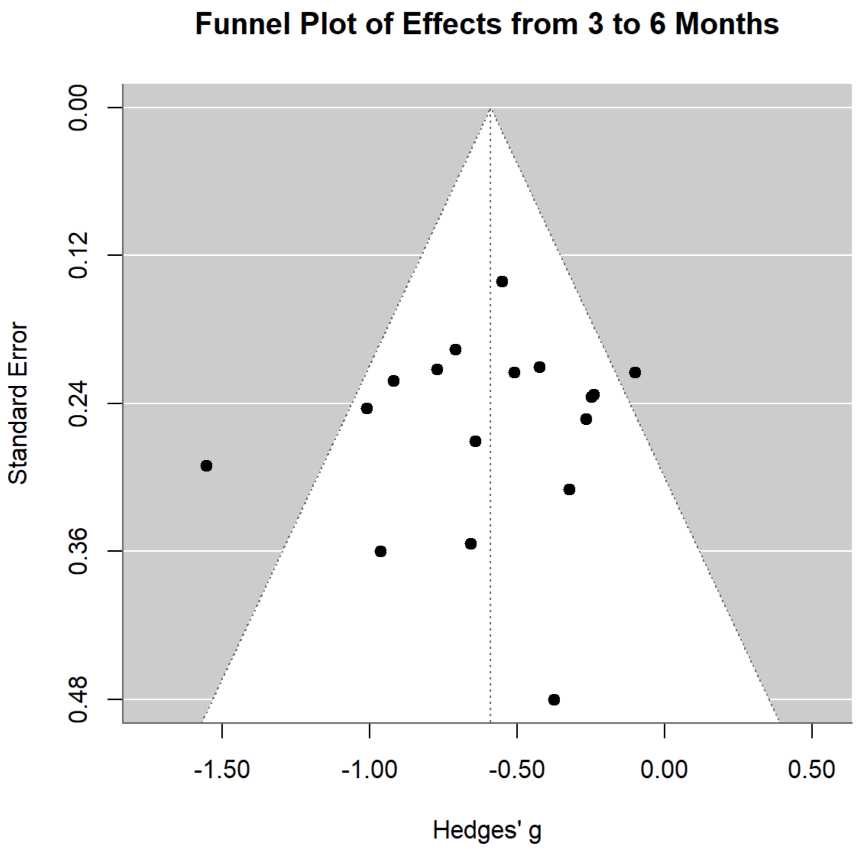


**Supplemental Figure 6.** The funnel plot for effects measured in the medium-term (from 3 months post-intervention to less than 6 months post-intervention).

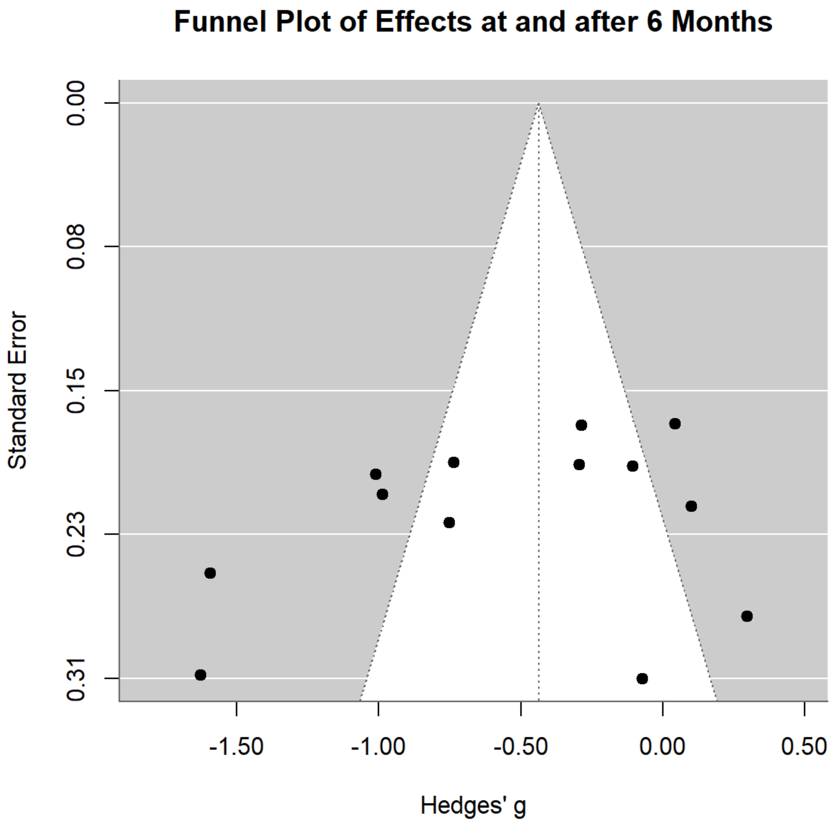


**Supplemental Figure 7.** The funnel plot for effects measured in the long-term (6 months or more post-intervention).

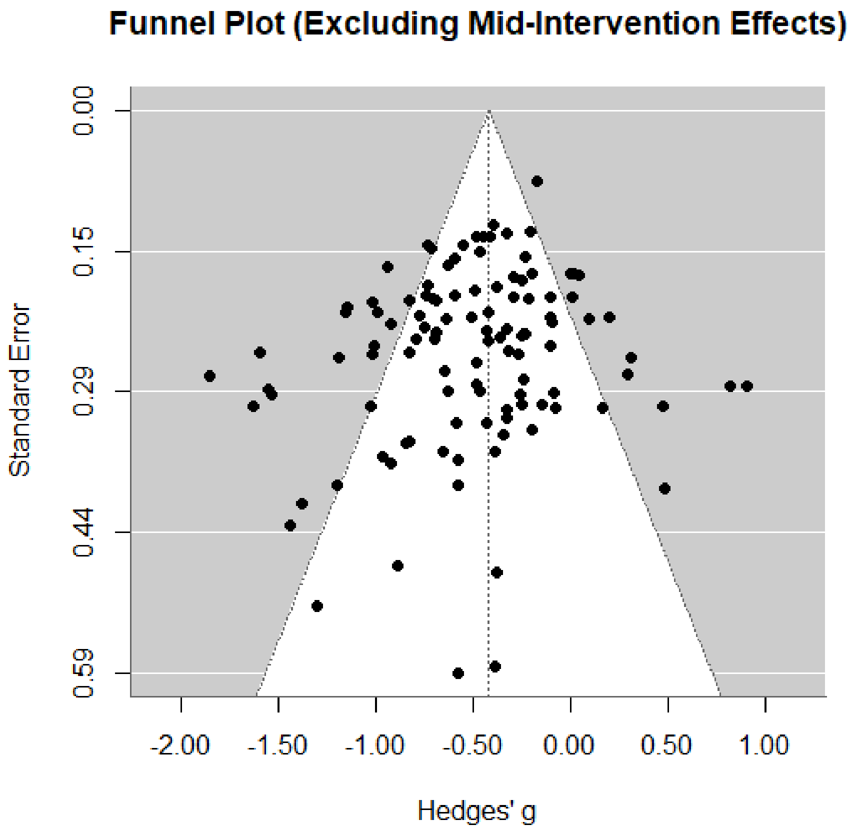


**Supplemental Figure 8.** The funnel plot excluding mid-intervention effects

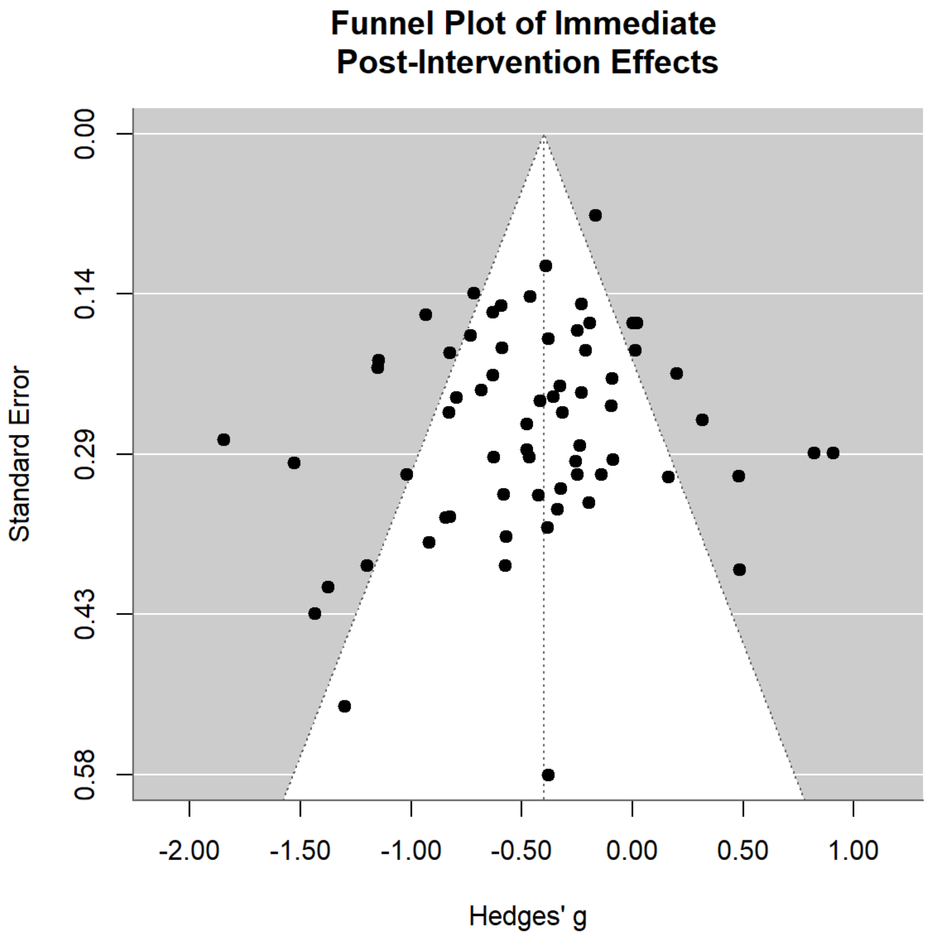


**Supplemental Figure 9.** The funnel plot for effects measured immediately post-intervention.

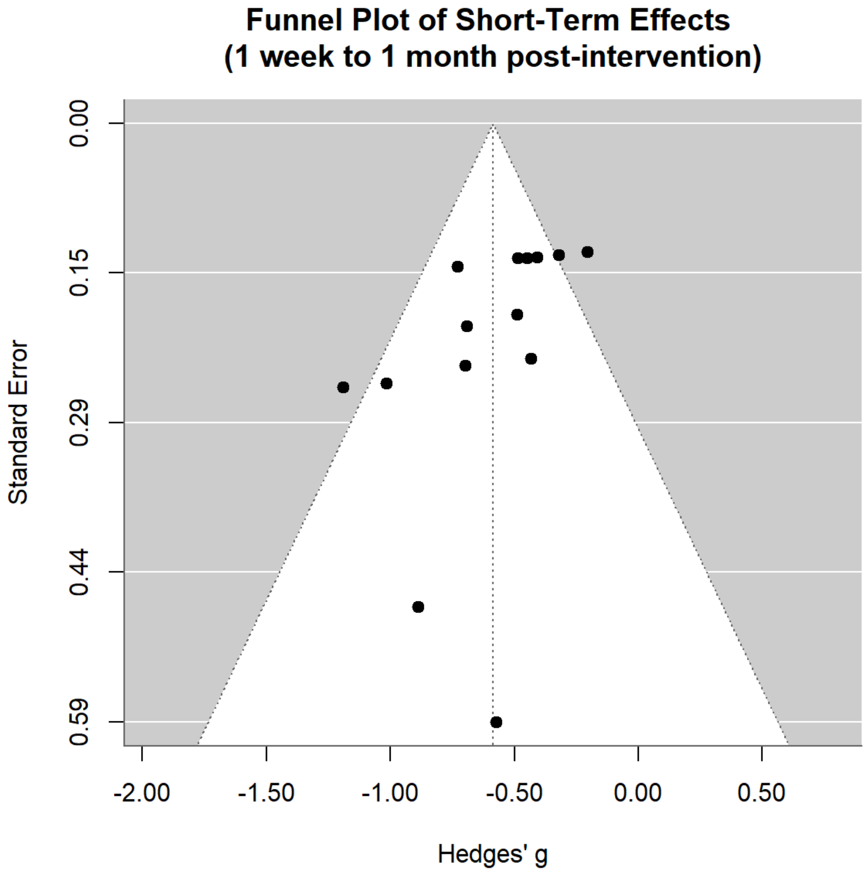


**Supplemental Figure 10.** The funnel plot for effects measured in the short-term, excluding those immediately post-intervention.

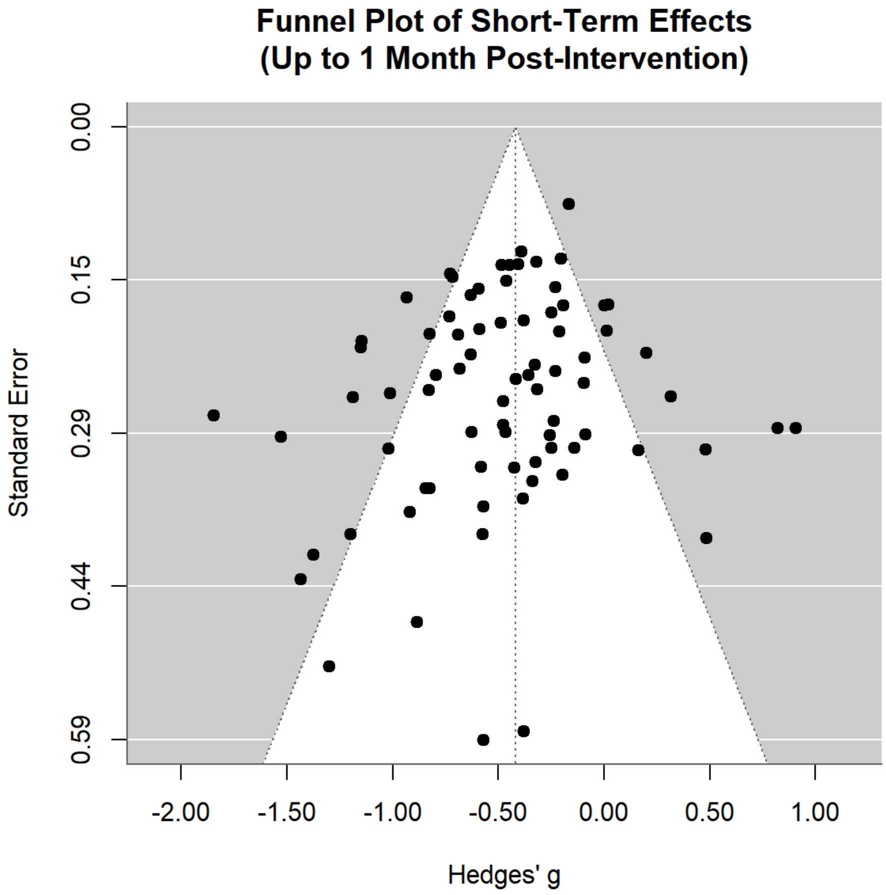


**Supplemental Figure 11.** The funnel plot for effects measured in the short-term, excluding those 2 months post-intervention.

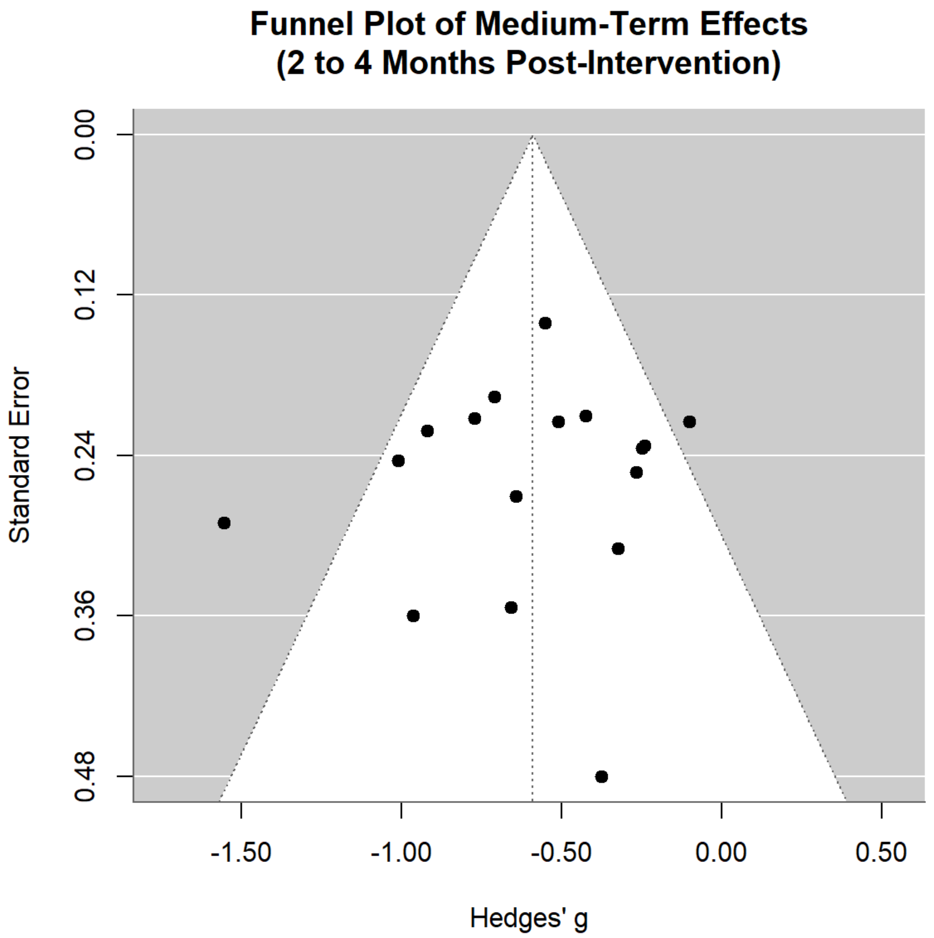


**Supplemental Figure 12.** The funnel plot for effects measured in the medium-term, including those 2 months post-intervention.
